# Impact of cannabis use on health outcomes and prescription benzodiazepine use

**DOI:** 10.1101/2025.08.23.25334295

**Authors:** Nirmal Singh, Yi Dai, Samuel T. Wilkinson, Taeho Greg Rhee, Rajiv Radhakrishnan

## Abstract

**Objective:** With the rising misuse of benzodiazepine (BZD) and associated overdose deaths, cannabis has been touted as a potential substitute with proposed benefit of better health outcomes. This two-year retrospective analysis examined whether cannabis use among BZD users was associated with changes in outcomes of (1) all-cause mortality, (2) hospitalizations, (3) emergency department (ED) visits, and (4) whether it demonstrated BZD-sparing effects on prescription quantity over time.

**Methods:** Using data from Yale New Haven Health System, we conducted a retrospective, longitudinal cohort study among BZD users. Cannabis use was the primary exposure, with BZD users without cannabis exposure as controls. Using inverse probability of treatment weighting and propensity score matching techniques, cohorts were balanced at baseline adjusting for medical comorbidities, socioeconomic status and other clinical factors. Kaplan-Meier curves and Cox proportional hazard models were examined with four outcomes of interest.

**Results:** The sample included 1,026 patients with 60.3% females and mean age was 54.2. There was no significant effect of cannabis use on BZD quantity. Cannabis use was not significantly associated with all-cause mortality, hospitalization, or ED visits. In exploratory analysis, medical cannabis users had a lower risk of all-cause mortality but greater risk for hospitalizations among those aged < 50 years.

**Conclusion:** In this longitudinal cohort study, cannabis use was not significantly associated with all-cause mortality, hospitalization and ED visits, or benzodiazepine dose reduction. Our results do not support a benzodiazepine-sparing effect for cannabis use.

## Introduction

Cannabis use is rapidly increasing across all demographics in the United States (US). In the US, the increase is driven by the growing legalization of recreational and medical use in more states [1], and the cannabis use has far outpaced the research needed to fully understand its health impacts [2]. Among young adults, any cannabis use in the past 12 months has steadily increased from 30.6% in 2013 to 42.4% in 2021 [3]. This trend is particularly pronounced in middle adulthood with the use increased from 13.9% in 2013 to 23.2% in 2021 [4]. Notably, a large proportion of the total cannabis use, up to 27% according to survey studies, is for medical purpose [2]. Among the primary reasons for medical cannabis use is the growing clinical interest in anxiolysis and in turn its “sparing effect,” which involves reducing the use of benzodiazepines among misusers and associated adverse public health outcomes [5, 6].

Benzodiazepines (BZDs) remain useful in the treatment of alcohol withdrawal, and in rare instances, for the treatment of anxiety and related disorders (e.g., insomnia and seizures) under supervision. However, BZDs are subject to misuse and are associated with the risk of overdose. In 2015 and 2016, 13% of Americans reported using BZDs in the past year, with 17-20% of such use being classified as misuse [7, 8]. Similarly, total number of overdose deaths involving benzodiazepines in the US surged from 1,135 in 1999 to 10,964 in 2022 [9, 10]. Beyond misuse and overdose fatalities, BZDs have been linked to potential increase in emergency department visits and all-cause mortality [11–15]. In a recent systemic review and meta-analysis, BZD use was associated with increased mortality with a hazard ratio ranging from 1.2 to 1.7 [16]. Furthermore, emergency department visits involving BZDs had a 20% increased risk of more serious health outcomes [17]. Given these risks, exploring therapeutic options that reduce or substitute the need for BZDs, such as the potential use of medical cannabis for its “sparing effects,” has been proposed [18].

The evidence supporting the use of cannabis for the treatment of anxiety and as a possible substitute for BZDs remains preliminary [19, 20]. Most of the studies asked for opinions which had inherent limitations. For example, a cross-sectional survey found that BZDs were the second most commonly substituted drugs with cannabis, with self-identified medical marijuana users reporting higher odds of substitution [21]. On the contrary, a study examining association of medical cannabis and BZD use in Canadian population did not reveal a significant change in BZD use among cannabis users. Similarly, O’Connell et al, found no reduction in diazepam equivalents required for pain control from baseline to six months in patients using medical cannabis [22]. The body of literature supporting medical cannabis as a substitute for BZDs remains limited, and studies examining its association with anxiety yielded inconclusive or mixed results. These gaps highlight the need for more patient-level longitudinal studies to better understand the therapeutic potentials and limitations of cannabis in regard to BZD use.

In our two-year retrospective study, we aimed to examine the impact of cannabis use on health outcomes among BZD users. In particular, we followed up and examined the outcomes of all-cause mortality, emergency department (ED) visits, hospitalizations. We also explored whether cannabis use had BZD-sparing effects. The findings of this study are expected to provide valuable insights into how cannabis use may influence benzodiazepine-related public health outcomes.

## Methods

### Data source and the study sample

Yale New Haven Health System (YNHH), located in the southern Connecticut of US, is a leading integrated health system consisted of five teaching hospitals with a total of 2,681 licensed beds, and with 151,850 inpatient visits and 5.2 million outpatient encounters annually. The study was approved by Yale University’s institutional review board (IRB) (HIC# 2000034126).

Retrospective data for all patients from YNHH between January 2022 and December 2023 was acquired from Yale Joint Data Analytics Team (JDAT). We included all adult patients aged 18 or older at an index date of May 1, 2022, and we followed them up through December 31, 2023. Inclusion criteria for this study were: (1) at least two encounters after the index date; (2) patients with at least 60 days of follow-up; (3) a history of BZD use during the pre-index period (i.e., January 1, 2022, through April 30, 2022) (**Figure 1**).

**Figure 1.**
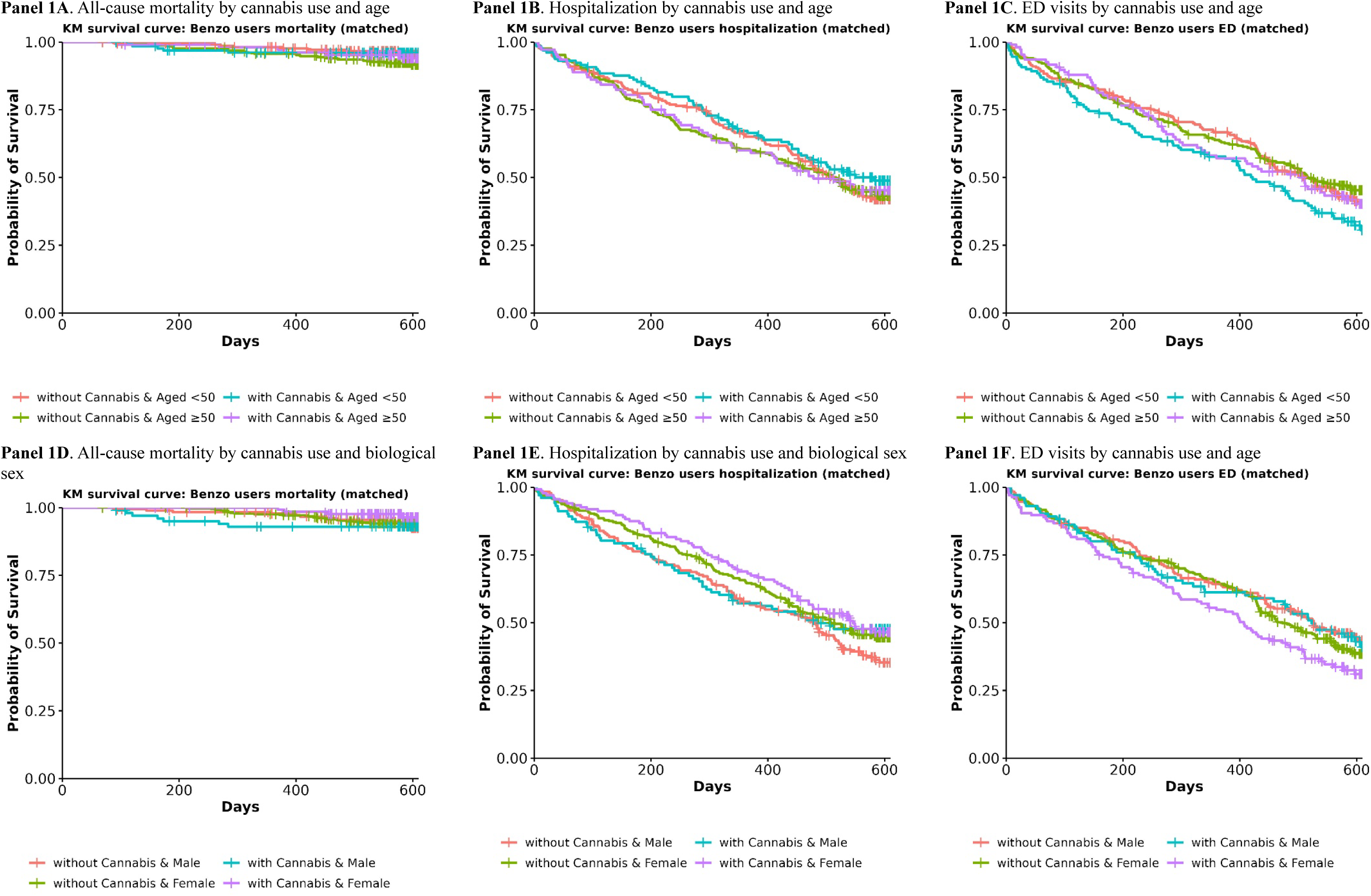
Kaplan-Meier survival curves for all-cause mortality, hospitalization, and emergency department (ED) visits by cannabis use according to age and biological sex among benzodiazepine users after propensity score matching. **Note:** Data are from 2022-2023 Yale-New Haven Health (YNHH).

### Measures

#### Cannabis use

The primary exposure measure was cannabis use (captured as yes or no). This measure was obtained from the Epic® database from YNHH and confirmed by urine toxicology positive for cannabinoids.

#### Outcome measures

Three dichotomous outcomes of interest included: all-cause mortality, hospitalization, and emergency department (ED) visits. All of these measures were ascertained through the Epic® database from YNHH.

#### Socio-demographic and clinical covariates

Other controlling variables included age, biological sex, race and ethnicity, insurance coverage (i.e., Medicaid, Medicare, private, or other), marital status (i.e., married, divorced or separated, widowed, or other), zip-code-level median income level, primary language use (i.e., English, Spanish, or other), and body mass index (BMI). All aforementioned variables were captured from the Epic® database in YNHH, except median income level, which were retrieved and linked with data from the 2024 American Community Survey (ACS) [23].

For clinical covariates, we included the medical modified (i.e., excluding psychiatric conditions) co-morbidities using Charleson Comorbidity Index (CCI) [24]. For psychiatric conditions, we included the following 10 categories: depression, anxiety, other mood disorders, psychosis, suicidal behaviors, alcohol use disorder, cannabis use disorder, opioid use disorder, other substance use disorders, or other. These clinical variables were based on the clinical diagnoses in the Epic® database from YNHH.

### Adjustment for confounding sources

Among patients with a history of BZD use, those engaged in cannabis use may have been inherently different from those who did not. Such differences may be a source of selection bias and may confound the true estimates of longitudinal health outcomes. To address such confounding factors, we balanced the characteristics of the two comparison groups (i.e., cannabis users and non-cannabis users) using all of observed socio-demographic and clinical characteristics between these two groups in the baseline (or pre-index) period (i.e., January 1, 2022, to April 30, 2022). Our primary approach was the propensity score matching (PSM) with a ratio of 1:2 (cannabis users to non-cannabis users) [25]. More specifically, we predicted the propensity scores for two comparison groups using socio-demographic and clinical covariates in the logistic regression model. Using the greedy nearest neighbor matching algorithm without replacement, each cannabis user (as a treated unit) was assigned to two non-cannabis users (as control units), who had the smallest distances based on the propensity scores. As part of the sensitivity analysis, we also performed inverse probability of treatment weighting (IPTW) to make the two comparison groups balanced at baseline [26]. We used analysis of variance (ANOVA) and chi-squared tests to test our covariate adjustments and balances at baseline.

### Analytical plan

We first described the socio-demographic and clinical characteristics by cannabis use at baseline (i.e., prior to the index date, May 1, 2022). Second, we followed up the study samples by cannabis use from the index date (i.e., May 1, 2022) throughout December 31, 2023. We estimated Kaplan-Meier curves to compare the survival probabilities of three main outcomes (i.e., all-cause mortality, hospitalization, and ED visits) between cannabis users and non-cannabis users among the whole study sample, and stratified by age and biological sex, respectively.

Then, we ran the Cox proportional hazards models handling ties with Efron’s method, to examine the conditional association of cannabis use (yes or no) and the survival time[27, 28]. Patients without the event of interests were censored at the date of the last encounter within the follow-up period. We estimated hazard ratios (HRs) with 95% confidence intervals, with a two-sided *p*<0.05 as the threshold of statistical significance.

The main analyses were those with PSM and we also reported findings with the adjustment of IPTW as part of the sensitivity analysis. As part of the sub-group analyses, we have also conducted 3-level exposure of cannabis use (i.e., non-user, cannabis use for medical purposes, and cannabis use for non-medical purposes). We also explored interaction effects by age and biological sex, respectively. We reported missing data patterns as part of the baseline patient characteristics, and missing data were handled using a listwise deletion approach in the multivariable-adjusted analyses. All analyses were conducted using R version 4.3.0., and the matching was conducted with a ‘*MatchIT*’ (version 4.5.5) package in R.

## Results

### Baseline characteristics of the study sample

The final analytic samples included 1,026 patients with a history of BZD use after meeting inclusion and exclusion criteria (**eFigure 1**), comprising of 238 (23.2%) who used cannabis and 788 (76.8%) non-cannabis users. On average, patients were 54.2 years old (standard deviation [SD], 15.9), 60.3% of patients were female and 68.8% of the patients were non-Hispanic white (**Table 1**). Most patients had either private (37.1%) or Medicaid (41.3%) insurance coverages, and the medial income level ranged from $50,000 to $89,999 (**Table 1**).

**Table 1.** Baseline socio-demographic and clinical characteristics of the study sample by cannabis use status before and after propensity score matching.

|  | Before matching |  |  |  | After matching (ratio 2:1) |  |  |  |
| --- | --- | --- | --- | --- | --- | --- | --- | --- |
|  | Non-user (n=788) | User (n=238) | Total (n=1,026) | P-value | Non-user (n=476) | User (n=238) | Total (n=714) | P-value |
| Age, mean ± standard deviation (SD) | 56.2 ± 15.9 | 47.4 ± 13.6 | 54.2 ± 15.8 | < 0.001 | 49.8 ± 15.0 | 47.4 ± 13.6 | 49.0 ± 14.6 | 0.037 |
| Biological sex |  |  |  |  |  |  |  |  |
| Male | 305 (38.7%) | 102 (42.9%) | 407 (39.7%) | 0.251 | 193 (40.5%) | 102 (42.9%) | 295 (41.3%) | 0.554 |
| Female | 483 (61.3%) | 136 (57.1%) | 619 (60.3%) |  | 283 (59.5%) | 136 (57.1%) | 419 (58.7%) |  |
| Race and ethnicity |  |  |  |  |  |  |  |  |
| Non-Hispanic white | 561 (71.2%) | 145 (60.9%) | 706 (68.8%) | <0.001 | 308 (64.7%) | 145 (60.9%) | 453 (63.4%) | 0.616 |
| Non-Hispanic black | 93 (11.8%) | 48 (20.2%) | 141 (13.7%) |  | 78 (16.4%) | 48 (20.2%) | 126 (17.6%) |  |
| Hispanic | 100 (12.7%) | 43 (18.1%) | 143 (13.9%) |  | 84 (17.6%) | 43 (18.1%) | 127 (17.8%) |  |
| Non-Hispanic other | 15 (1.9%) | 0 (0.0%) | 15 (1.5%) |  | 2 (0.4%) | 0 (0.0%) | 2 (0.3%) |  |
| Unknown/missing | 19 (2.4%) | 2 (0.8%) | 21 (2.0%) |  | 4 (0.8%) | 2 (0.8%) | 6 (0.8%) |  |
| Marital status |  |  |  |  |  |  |  |  |
| Single | 328 (41.6%) | 137 (57.6%) | 465 (45.3%) | <0.001 | 259 (54.4%) | 137 (57.6%) | 396 (55.5%) | 0.913 |
| Married | 245 (31.1%) | 43 (18.1%) | 288 (28.1%) |  | 88 (18.5%) | 43 (18.1%) | 131 (18.3%) |  |
| Divorced or separated | 128 (16.2%) | 35 (14.7%) | 163 (15.9%) |  | 78 (16.4%) | 35 (14.7%) | 113 (15.8%) |  |
| Widowed | 57 (7.2%) | 14 (5.9%) | 71 (6.9%) |  | 34 (7.1%) | 14 (5.9%) | 48 (6.7%) |  |
| Other | 30 (3.8%) | 9 (3.8%) | 39 (3.8%) |  | 17 (3.6%) | 9 (3.8%) | 26 (3.6%) |  |
| Income level |  |  |  |  |  |  |  |  |
| <50,000 | 116 (14.7%) | 41 (17.2%) | 157 (15.3%) | 0.056 | 85 (17.9%) | 41 (17.2%) | 126 (17.6%) | 0.825 |
| 50,000-89,999 | 317 (40.2%) | 116 (48.7%) | 433 (42.2%) |  | 221 (46.4%) | 116 (48.7%) | 337 (47.2%) |  |
| 90,000-129,999 | 273 (34.6%) | 63 (26.5%) | 336 (32.7%) |  | 139 (29.2%) | 63 (26.5%) | 202 (28.3%) |  |
| 130,000-149,999 | 38 (4.8%) | 9 (3.8%) | 47 (4.6%) |  | 19 (4.0%) | 9 (3.8%) | 28 (3.9%) |  |
| ≥150,000 | 44 (5.6%) | 9 (3.8%) | 53 (5.2%) | | 12 (2.5%) | 9 (3.8%) | 21 (2.9%) | |
| Insurance coverage |  |  |  |  |  |  |  |  |
| Private | 311 (39.5%) | 70 (29.4%) | 381 (37.1%) | <0.001 | 150 (31.5%) | 70 (29.4%) | 220 (30.8%) | 0.983 |
| Medicaid | 291 (36.9%) | 133 (55.9%) | 424 (41.3%) |  | 258 (54.2%) | 133 (55.9%) | 391 (54.8%) |  |
| Medicare | 145 (18.4%) | 23 (9.7%) | 168 (16.4%) |  | 43 (9.0%) | 23 (9.7%) | 66 (9.2%) |  |
| Other | 31 (3.9%) | 8 (3.4%) | 39 (3.8%) |  | 17 (3.6%) | 8 (3.4%) | 25 (3.5%) |  |
| Unknown/missing | 10 (1.3%) | 4 (1.7%) | 14 (1.4%) |  | 8 (1.7%) | 4 (1.7%) | 12 (1.7%) |  |
| Primary language use |  |  |  |  |  |  |  |  |
| English | 768 (97.5%) | 234 (98.3%) | 1002 (97.7%) | 0.745 | 464 (97.5%) | 234 (98.3%) | 698 (97.8%) | 0.561 |
| Spanish | 15 (1.9%) | 3 (1.3%) | 18 (1.8%) |  | 11 (2.3%) | 3 (1.3%) | 14 (2.0%) |  |
| Other | 5 (0.6%) | 1 (0.4%) | 6 (0.6%) |  | 1 (0.2%) | 1 (0.4%) | 2 (0.3%) |  |
| BMI, mean ± SD | 29.4 ± 8.0 | 29.3 ± 7.8 | 29.4 ± 7.9 | 0.842 | 29.9 ± 8.4 | 29.3 ± 7.8 | 29.7 ± 8.2 | 0.379 |
| Charlson comorbidity index |  |  |  |  |  |  |  |  |
| None | 352 (44.7%) | 117 (49.2%) | 469 (45.7%) | 0.544 | 230 (48.3%) | 117 (49.2%) | 347 (48.6%) | 0.759 |
| 1-2 | 235 (29.8%) | 68 (28.6%) | 303 (29.5%) |  | 131 (27.5%) | 68 (28.6%) | 199 (27.9%) |  |
| 3-4 | 99 (12.6%) | 29 (12.2%) | 128 (12.5%) |  | 54 (11.3%) | 29 (12.2%) | 83 (11.6%) |  |
| ≥5 | 102 (12.9%) | 24 (10.1%) | 126 (12.3%) | | 61 (12.8%) | 24 (10.1%) | 85 (11.9%) | |
| Current alcohol use |  |  |  |  |  |  |  |  |
| No | 297 (37.7%) | 81 (34.0%) | 378 (36.8%) | 0.605 | 176 (37.0%) | 81 (34.0%) | 257 (36.0%) | 0.766 |
| Yes | 241 (30.6%) | 82 (34.5%) | 323 (31.5%) |  | 147 (30.9%) | 82 (34.5%) | 229 (32.1%) |  |
| Not currently | 244 (31.0%) | 74 (31.1%) | 318 (31.0%) |  | 150 (31.5%) | 74 (31.1%) | 224 (31.4%) |  |
| Unknown/missing | 6 (0.8%) | 1 (0.4%) | 7 (0.7%) |  | 3 (0.6%) | 1 (0.4%) | 4 (0.6%) |  |
| Current tobacco use |  |  |  |  |  |  |  |  |
| Never | 290 (36.8%) | 52 (21.8%) | 342 (33.3%) |  | 173 (36.3%) | 52 (21.8%) | 225 (31.5%) |  |
| Yes | 194 (24.6%) | 99 (41.6%) | 293 (28.6%) | < 0.001 | 148 (31.1%) | 99 (41.6%) | 247 (34.6%) | <0.001 |
| Quitted | 303 (38.5%) | 87 (36.6%) | 390 (38.0%) |  | 154 (32.4%) | 87 (36.6%) | 241 (33.8%) |  |
| Other | 1 (0.1%) | 0 (0.0%) | 1 (0.1%) |  | 1 (0.2%) | 0 (0.0%) | 1 (0.1%) |  |
| Depression |  |  |  |  |  |  |  |  |
| No | 224 (28.4%) | 71 (29.8%) | 295 (28.8%) |  | 138 (29.0%) | 71 (29.8%) | 209 (29.3%) |  |
| Yes | 564 (71.6%) | 167 (70.2%) | 731 (71.2%) | 0.675 | 338 (71.0%) | 167 (70.2%) | 505 (70.7%) | 0.816 |
| Anxiety |  |  |  |  |  |  |  |  |
| No | 618 (78.4%) | 193 (81.1%) | 811 (79.0%) |  | 379 (79.6%) | 193 (81.1%) | 572 (80.1%) |  |
| Yes | 170 (21.6%) | 45 (18.9%) | 215 (21.0%) | 0.376 | 97 (20.4%) | 45 (18.9%) | 142 (19.9%) | 0.643 |
| Other mood disorder |  |  |  |  |  |  |  |  |
| No | 583 (74.0%) | 152 (63.9%) | 735 (71.6%) |  | 325 (68.3%) | 152 (63.9%) | 477 (66.8%) |  |
| Yes | 205 (26.0%) | 86 (36.1%) | 291 (28.4%) | 0.002 | 151 (31.7%) | 86 (36.1%) | 237 (33.2%) | 0.238 |
| Suicidal symptoms |  |  |  |  |  |  |  |  |
| No | 741 (94.0%) | 224 (94.1%) | 965 (94.1%) |  | 443 (93.1%) | 224 (94.1%) | 667 (93.4%) |  |
| Yes | 47 (6.0%) | 14 (5.9%) | 61 (5.9%) | 0.963 | 33 (6.9%) | 14 (5.9%) | 47 (6.6%) | 0.594 |
| Psychosis |  |  |  |  |  |  |  |  |
| No | 693 (87.9%) | 208 (87.4%) | 901 (87.8%) |  | 422 (88.7%) | 208 (87.4%) | 630 (88.2%) |  |
| Yes | 95 (12.1%) | 30 (12.6%) | 125 (12.2%) | 0.820 | 54 (11.3%) | 30 (12.6%) | 84 (11.8%) | 0.622 |
| Alcohol use disorder |  |  |  |  |  |  |  |  |
| No | 728 (92.4%) | 219 (92.0%) | 947 (92.3%) |  | 439 (92.2%) | 219 (92.0%) | 658 (92.2%) |  |
| Yes | 60 (7.6%) | 19 (8.0%) | 79 (7.7%) | 0.852 | 37 (7.8%) | 19 (8.0%) | 56 (7.8%) | 0.922 |
| Opioid use disorder |  |  |  |  |  |  |  |  |
| No | 732 (92.9%) | 220 (92.4%) | 952 (92.8%) |  | 445 (93.5%) | 220 (92.4%) | 665 (93.1%) |  |
| Yes | 56 (7.1%) | 18 (7.6%) | 74 (7.2%) | 0.811 | 31 (6.5%) | 18 (7.6%) | 49 (6.9%) | 0.601 |
| Cannabis use disorder |  |  |  |  |  |  |  |  |
| No | 779 (98.9%) | 217 (91.2%) | 996 (97.1%) |  | 467 (98.1%) | 217 (91.2%) | 684 (95.8%) |  |
| Yes | 9 (1.1%) | 21 (8.8%) | 30 (2.9%) | <0.001 | 9 (1.9%) | 21 (8.8%) | 30 (4.2%) | <0.001 |
| Other substance use disorder |  |  |  |  |  |  |  |  |
| No | 720 (91.4%) | 204 (85.7%) | 924 (90.1%) |  | 422 (88.7%) | 204 (85.7%) | 626 (87.7%) |  |
| Yes | 68 (8.6%) | 34 (14.3%) | 102 (9.9%) | 0.011 | 54 (11.3%) | 34 (14.3%) | 88 (12.3%) | 0.260 |
| Other psychiatric condition |  |  |  |  |  |  |  |  |
| No | 767 (97.3%) | 228 (95.8%) | 995 (97.0%) |  | 459 (96.4%) | 228 (95.8%) | 687 (96.2%) |  |
| Yes | 21 (2.7%) | 10 (4.2%) | 31 (3.0%) | 0.225 | 17 (3.6%) | 10 (4.2%) | 27 (3.8%) | 0.677 |

**Note:** Data are from 2022-2023 Yale-New Haven Health (YNHH).

At baseline, the distributions of several variables were significantly different between cannabis users and non-cannabis users. When the PSM was applied, most of the variables became not significantly different, except three variables: age, current tobacco use, and cannabis use disorder-these were included as covariates in the analysis. Using the IPTW approach, we found similar distributions of variables at baseline (**Table S1**), except three variables that had significantly different distributions: race/ethnicity, income level, and current tobacco use-these were included as covariates in the analysis. Overall, most variables were similar across two different comparison groups at baseline.

In our raw sample, rate of all-cause mortality was 0.07±0.26 % (Cannabis 0.05±0.21 %, Non-cannabis 0.08±0.27 %), ED visits was 0.53±0.26 % (Cannabis 0.60 ± 0.21 %, Non-cannabis 0.57±0.27 %), and hospitalization was 0.54±0.26 % (Cannabis 0.51 ± 0.21 %, Non-cannabis 0.55±0.27 %).

The average baseline benzodiazepine dose was 8.13 diazepam milligram equivalency (DME) [29], and the median was 3.13 DME; there were no baseline group differences in DME. The average [and median] follow-up days until the end of the study were similar across comparison groups (i.e., 408.5 [444.5] days for non-cannabis users and 411.2 [453.0] days for cannabis users). The mean [and median] numbers of encounters were also similar across groups (i.e., 44.2 [30.0] encounters for non-cannabis users and 40.7 [26.5] encounters for cannabis users).

### Associations of cannabis use with outcomes of interest: Main analyses

Unadjusted Kaplan-Meier curves are displayed in **Figure 1**, which were stratified by age (i.e., **Panels 1A to 1C**) and biological sex (i.e., **Panels 1D to 1F**), respectively. We also reported the same analyses using the IPTW approach in **Figure S2**. In the main multivariable-adjusted Cox proportional hazards models (**Table 2**), we did not find any statistical difference in the potential impacts of cannabis use on all-cause mortality, hospitalization, and ED visits, respectively.

**Table 2.**
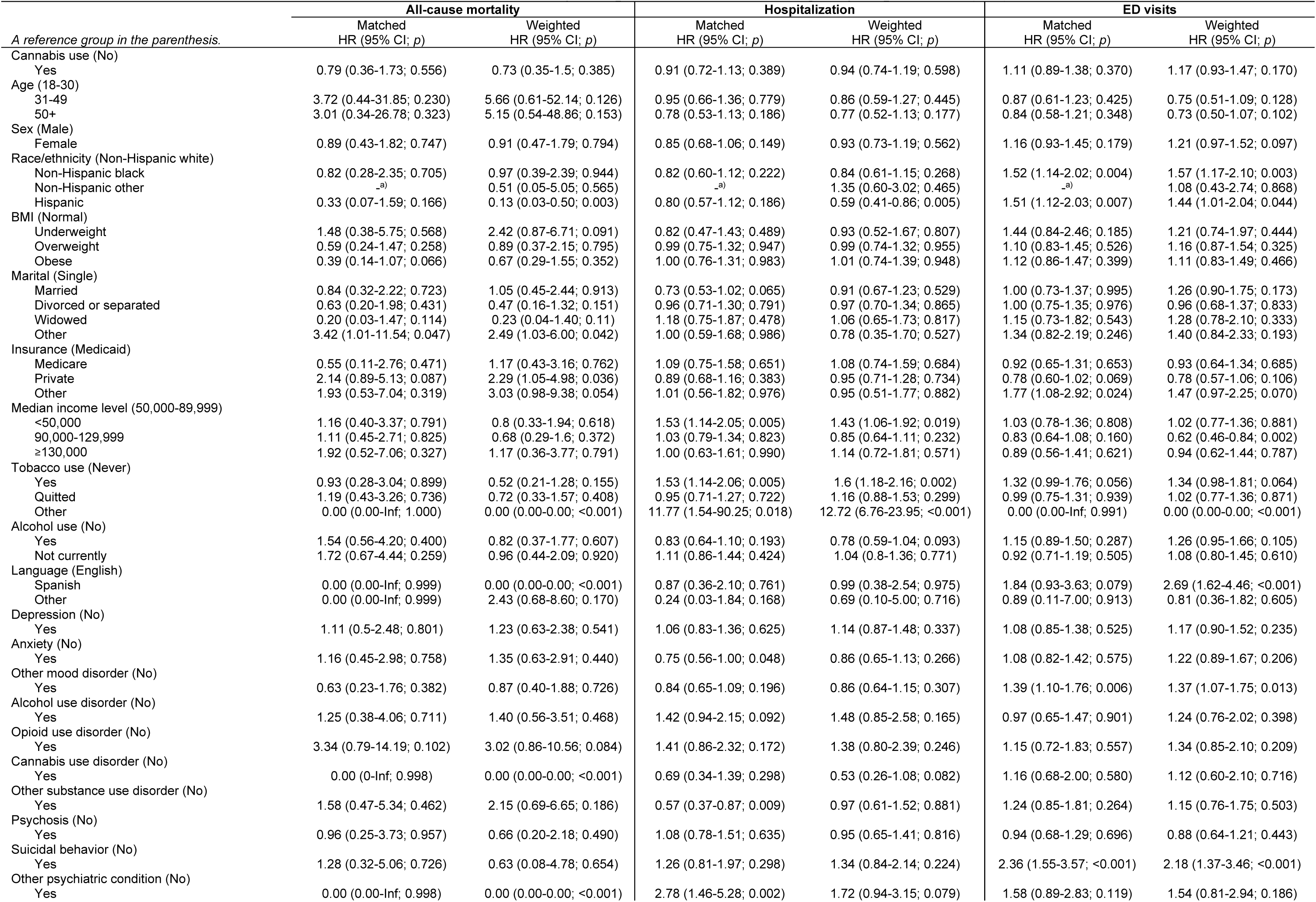
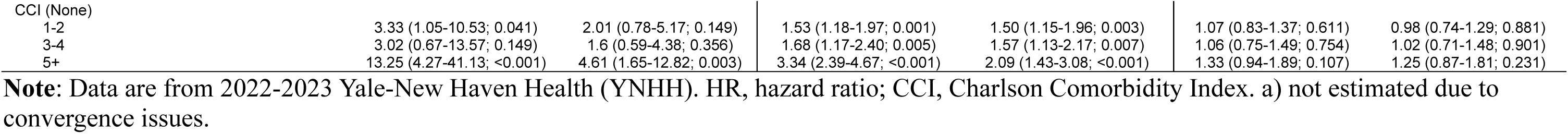
Cannabis use and its association with all-cause mortality, hospitalization, and emergency department (ED) visits.

| *A reference group in the parenthesis.* | All-cause mortality |  | Hospitalization |  | ED visits |  |
| --- | --- | --- | --- | --- | --- | --- |
|  | Matched<br>HR (95% CI; *p*) | Weighted<br>HR (95% CI; *p*) | Matched<br>HR (95% CI; *p*) | Weighted<br>HR (95% CI; *p*) | Matched<br>HR (95% CI; *p*) | Weighted<br>HR (95% CI; *p*) |
| Cannabis use (No) |  |  |  |  |  |  |
| Yes | 0.79 (0.36-1.73; 0.556) | 0.73 (0.35-1.5; 0.385) | 0.91 (0.72-1.13; 0.389) | 0.94 (0.74-1.19; 0.598) | 1.11 (0.89-1.38; 0.370) | 1.17 (0.93-1.47; 0.170) |
| Age (18-30) |  |  |  |  |  |  |
| 31-49 | 3.72 (0.44-31.85; 0.230) | 5.66 (0.61-52.14; 0.126) | 0.95 (0.66-1.36; 0.779) | 0.86 (0.59-1.27; 0.445) | 0.87 (0.61-1.23; 0.425) | 0.75 (0.51-1.09; 0.128) |
| 50+ | 3.01 (0.34-26.78; 0.323) | 5.15 (0.54-48.86; 0.153) | 0.78 (0.53-1.13; 0.186) | 0.77 (0.52-1.13; 0.177) | 0.84 (0.58-1.21; 0.348) | 0.73 (0.50-1.07; 0.102) |
| Sex (Male) |  |  |  |  |  |  |
| Female | 0.89 (0.43-1.82; 0.747) | 0.91 (0.47-1.79; 0.794) | 0.85 (0.68-1.06; 0.149) | 0.93 (0.73-1.19; 0.562) | 1.16 (0.93-1.45; 0.179) | 1.21 (0.97-1.52; 0.097) |
| Race/ethnicity (Non-Hispanic white) |  |  |  |  |  |  |
| Non-Hispanic black | 0.82 (0.28-2.35; 0.705) | 0.97 (0.39-2.39; 0.944) | 0.82 (0.60-1.12; 0.222) | 0.84 (0.61-1.15; 0.268) | 1.52 (1.14-2.02; 0.004) | 1.57 (1.17-2.10; 0.003) |
| Non-Hispanic other | <sup>a)</sup> | 0.51 (0.05-5.05; 0.565) | <sup>a)</sup> | 1.35 (0.60-3.02; 0.465) | <sup>a)</sup> | 1.08 (0.43-2.74; 0.868) |
| Hispanic | 0.33 (0.07-1.59; 0.166) | 0.13 (0.03-0.50; 0.003) | 0.80 (0.57-1.12; 0.186) | 0.59 (0.41-0.86; 0.005) | 1.51 (1.12-2.03; 0.007) | 1.44 (1.01-2.04; 0.044) |
| BMI (Normal) |  |  |  |  |  |  |
| Underweight | 1.48 (0.38-5.75; 0.568) | 2.42 (0.87-6.71; 0.091) | 0.82 (0.47-1.43; 0.489) | 0.93 (0.52-1.67; 0.807) | 1.44 (0.84-2.46; 0.185) | 1.21 (0.74-1.97; 0.444) |
| Overweight | 0.59 (0.24-1.47; 0.258) | 0.89 (0.37-2.15; 0.795) | 0.99 (0.75-1.32; 0.947) | 0.99 (0.74-1.32; 0.955) | 1.10 (0.83-1.45; 0.526) | 1.16 (0.87-1.54; 0.325) |
| Obese | 0.39 (0.14-1.07; 0.066) | 0.67 (0.29-1.55; 0.352) | 1.00 (0.76-1.31; 0.983) | 1.01 (0.74-1.39; 0.948) | 1.12 (0.86-1.47; 0.399) | 1.11 (0.83-1.49; 0.466) |
| Marital (Single) |  |  |  |  |  |  |
| Married | 0.84 (0.32-2.22; 0.723) | 1.05 (0.45-2.44; 0.913) | 0.73 (0.53-1.02; 0.065) | 0.91 (0.67-1.23; 0.529) | 1.00 (0.73-1.37; 0.995) | 1.26 (0.90-1.75; 0.173) |
| Divorced or separated | 0.63 (0.20-1.98; 0.431) | 0.47 (0.16-1.32; 0.151) | 0.96 (0.71-1.30; 0.791) | 0.97 (0.70-1.34; 0.865) | 1.00 (0.75-1.35; 0.976) | 0.96 (0.68-1.37; 0.833) |
| Widowed | 0.20 (0.03-1.47; 0.114) | 0.23 (0.04-1.40; 0.11) | 1.18 (0.75-1.87; 0.478) | 1.06 (0.65-1.73; 0.817) | 1.15 (0.73-1.82; 0.543) | 1.28 (0.78-2.10; 0.333) |
| Other | 3.42 (1.01-11.54; 0.047) | 2.49 (1.03-6.00; 0.042) | 1.00 (0.59-1.68; 0.986) | 0.78 (0.35-1.70; 0.527) | 1.34 (0.82-2.19; 0.246) | 1.40 (0.84-2.33; 0.193) |
| Insurance (Medicaid) |  |  |  |  |  |  |
| Medicare | 0.55 (0.11-2.76; 0.471) | 1.17 (0.43-3.16; 0.762) | 1.09 (0.75-1.58; 0.651) | 1.08 (0.74-1.59; 0.684) | 0.92 (0.65-1.31; 0.653) | 0.93 (0.64-1.34; 0.685) |
| Private | 2.14 (0.89-5.13; 0.087) | 2.29 (1.05-4.98; 0.036) | 0.89 (0.68-1.16; 0.383) | 0.95 (0.71-1.28; 0.734) | 0.78 (0.60-1.02; 0.069) | 0.78 (0.57-1.06; 0.106) |
| Other | 1.93 (0.53-7.04; 0.319) | 3.03 (0.98-9.38; 0.054) | 1.01 (0.56-1.82; 0.976) | 0.95 (0.51-1.77; 0.882) | 1.77 (1.08-2.92; 0.024) | 1.47 (0.97-2.25; 0.070) |
| Median income level (50,000-89,999) |  |  |  |  |  |  |
| <50,000 | 1.16 (0.40-3.37; 0.791) | 0.8 (0.33-1.94; 0.618) | 1.53 (1.14-2.05; 0.005) | 1.43 (1.06-1.92; 0.019) | 1.03 (0.78-1.36; 0.808) | 1.02 (0.77-1.36; 0.881) |
| 90,000-129,999 | 1.11 (0.45-2.71; 0.825) | 0.68 (0.29-1.6; 0.372) | 1.03 (0.79-1.34; 0.823) | 0.85 (0.64-1.11; 0.232) | 0.83 (0.64-1.08; 0.160) | 0.62 (0.46-0.84; 0.002) |
| ≥130,000 | 1.92 (0.52-7.06; 0.327) | 1.17 (0.36-3.77; 0.791) | 1.00 (0.63-1.61; 0.990) | 1.14 (0.72-1.81; 0.571) | 0.89 (0.56-1.41; 0.621) | 0.94 (0.62-1.44; 0.787) |
| Tobacco use (Never) |  |  |  |  |  |  |
| Yes | 0.93 (0.28-3.04; 0.899) | 0.52 (0.21-1.28; 0.155) | 1.53 (1.14-2.06; 0.005) | 1.6 (1.18-2.16; 0.002) | 1.32 (0.99-1.76; 0.056) | 1.34 (0.98-1.81; 0.064) |
| Quitted | 1.19 (0.43-3.26; 0.736) | 0.72 (0.33-1.57; 0.408) | 0.95 (0.71-1.27; 0.722) | 1.16 (0.88-1.53; 0.299) | 0.99 (0.75-1.31; 0.939) | 1.02 (0.77-1.36; 0.871) |
| Other | 0.00 (0.00-Inf; 1.000) | 0.00 (0.00-0.00; <0.001) | 11.77 (1.54-90.25; 0.018) | 12.72 (6.76-23.95; <0.001) | 0.00 (0.00-Inf; 0.991) | 0.00 (0.00-0.00; <0.001) |
| Alcohol use (No) |  |  |  |  |  |  |
| Yes | 1.54 (0.56-4.20; 0.400) | 0.82 (0.37-1.77; 0.607) | 0.83 (0.64-1.10; 0.193) | 0.78 (0.59-1.04; 0.093) | 1.15 (0.89-1.50; 0.287) | 1.26 (0.95-1.66; 0.105) |
| Not currently | 1.72 (0.67-4.44; 0.259) | 0.96 (0.44-2.09; 0.920) | 1.11 (0.86-1.44; 0.424) | 1.04 (0.8-1.36; 0.771) | 0.92 (0.71-1.19; 0.505) | 1.08 (0.80-1.45; 0.610) |
| Language (English) |  |  |  |  |  |  |
| Spanish | 0.00 (0.00-Inf; 0.999) | 0.00 (0.00-0.00; <0.001) | 0.87 (0.36-2.10; 0.761) | 0.99 (0.38-2.54; 0.975) | 1.84 (0.93-3.63; 0.079) | 2.69 (1.62-4.46; <0.001) |
| Other | 0.00 (0.00-Inf; 0.999) | 2.43 (0.68-8.60; 0.170) | 0.24 (0.03-1.84; 0.168) | 0.69 (0.10-5.00; 0.716) | 0.89 (0.11-7.00; 0.913) | 0.81 (0.36-1.82; 0.605) |
| Depression (No) |  |  |  |  |  |  |
| Yes | 1.11 (0.5-2.48; 0.801) | 1.23 (0.63-2.38; 0.541) | 1.06 (0.83-1.36; 0.625) | 1.14 (0.87-1.48; 0.337) | 1.08 (0.85-1.38; 0.525) | 1.17 (0.90-1.52; 0.235) |
| Anxiety (No) |  |  |  |  |  |  |
| Yes | 1.16 (0.45-2.98; 0.758) | 1.35 (0.63-2.91; 0.440) | 0.75 (0.56-1.00; 0.048) | 0.86 (0.65-1.13; 0.266) | 1.08 (0.82-1.42; 0.575) | 1.22 (0.89-1.67; 0.206) |
| Other mood disorder (No) |  |  |  |  |  |  |
| Yes | 0.63 (0.23-1.76; 0.382) | 0.87 (0.40-1.88; 0.726) | 0.84 (0.65-1.09; 0.196) | 0.86 (0.64-1.15; 0.307) | 1.39 (1.10-1.76; 0.006) | 1.37 (1.07-1.75; 0.013) |
| Alcohol use disorder (No) |  |  |  |  |  |  |
| Yes | 1.25 (0.38-4.06; 0.711) | 1.40 (0.56-3.51; 0.468) | 1.42 (0.94-2.15; 0.092) | 1.48 (0.85-2.58; 0.165) | 0.97 (0.65-1.47; 0.901) | 1.24 (0.76-2.02; 0.398) |
| Opioid use disorder (No) |  |  |  |  |  |  |
| Yes | 3.34 (0.79-14.19; 0.102) | 3.02 (0.86-10.56; 0.084) | 1.41 (0.86-2.32; 0.172) | 1.38 (0.80-2.39; 0.246) | 1.15 (0.72-1.83; 0.557) | 1.34 (0.85-2.10; 0.209) |
| Cannabis use disorder (No) |  |  |  |  |  |  |
| Yes | 0.00 (0-Inf; 0.998) | 0.00 (0.00-0.00; <0.001) | 0.69 (0.34-1.39; 0.298) | 0.53 (0.26-1.08; 0.082) | 1.16 (0.68-2.00; 0.580) | 1.12 (0.60-2.10; 0.716) |
| Other substance use disorder (No) |  |  |  |  |  |  |
| Yes | 1.58 (0.47-5.34; 0.462) | 2.15 (0.69-6.65; 0.186) | 0.57 (0.37-0.87; 0.009) | 0.97 (0.61-1.52; 0.881) | 1.24 (0.85-1.81; 0.264) | 1.15 (0.76-1.75; 0.503) |
| Psychosis (No) |  |  |  |  |  |  |
| Yes | 0.96 (0.25-3.73; 0.957) | 0.66 (0.20-2.18; 0.490) | 1.08 (0.78-1.51; 0.635) | 0.95 (0.65-1.41; 0.816) | 0.94 (0.68-1.29; 0.696) | 0.88 (0.64-1.21; 0.443) |
| Suicidal behavior (No) |  |  |  |  |  |  |
| Yes | 1.28 (0.32-5.06; 0.726) | 0.63 (0.08-4.78; 0.654) | 1.26 (0.81-1.97; 0.298) | 1.34 (0.84-2.14; 0.224) | 2.36 (1.55-3.57; <0.001) | 2.18 (1.37-3.46; <0.001) |
| Other psychiatric condition (No) |  |  |  |  |  |  |
| Yes | 0.00 (0.00-Inf; 0.998) | 0.00 (0.00-0.00; <0.001) | 2.78 (1.46-5.28; 0.002) | 1.72 (0.94-3.15; 0.079) | 1.58 (0.89-2.83; 0.119) | 1.54 (0.81-2.94; 0.186) |
| CCI (None) |  |  |  |  |  |  |
| 1-2 | 3.33 (1.05-10.53; 0.041) | 2.01 (0.78-5.17; 0.149) | 1.53 (1.18-1.97; 0.001) | 1.50 (1.15-1.96; 0.003) | 1.07 (0.83-1.37; 0.611) | 0.98 (0.74-1.29; 0.881) |
| 3-4 | 3.02 (0.67-13.57; 0.149) | 1.6 (0.59-4.38; 0.356) | 1.68 (1.17-2.40; 0.005) | 1.57 (1.13-2.17; 0.007) | 1.06 (0.75-1.49; 0.754) | 1.02 (0.71-1.48; 0.901) |
| 5+ | 13.25 (4.27-41.13; <0.001) | 4.61 (1.65-12.82; 0.003) | 3.34 (2.39-4.67; <0.001) | 2.09 (1.43-3.08; <0.001) | 1.33 (0.94-1.89; 0.107) | 1.25 (0.87-1.81; 0.231) |

**Note:** Data are from 2022-2023 Yale-New Haven Health (YNHH). HR, hazard ratio; CCI, Charlson Comorbidity Index. a) not estimated due to convergence issues.

When exploring for the potential interaction effects of cannabis use and age on the outcomes of interest (**Table 3**), we found that patients aged 50 or older and with non-cannabis use had a greater hazard rate of all-cause mortality (cumulative HR [cHR], 2.17 [95% CI, 1.00-4.69]; *p*=0.049) when compared to those aged 49 or younger and with non-cannabis use in the IPTW-adjusted model. We did not find any significant interaction effects of biological sex and cannabis use on the outcomes of interest (**Table 4**).

**Table 3.**
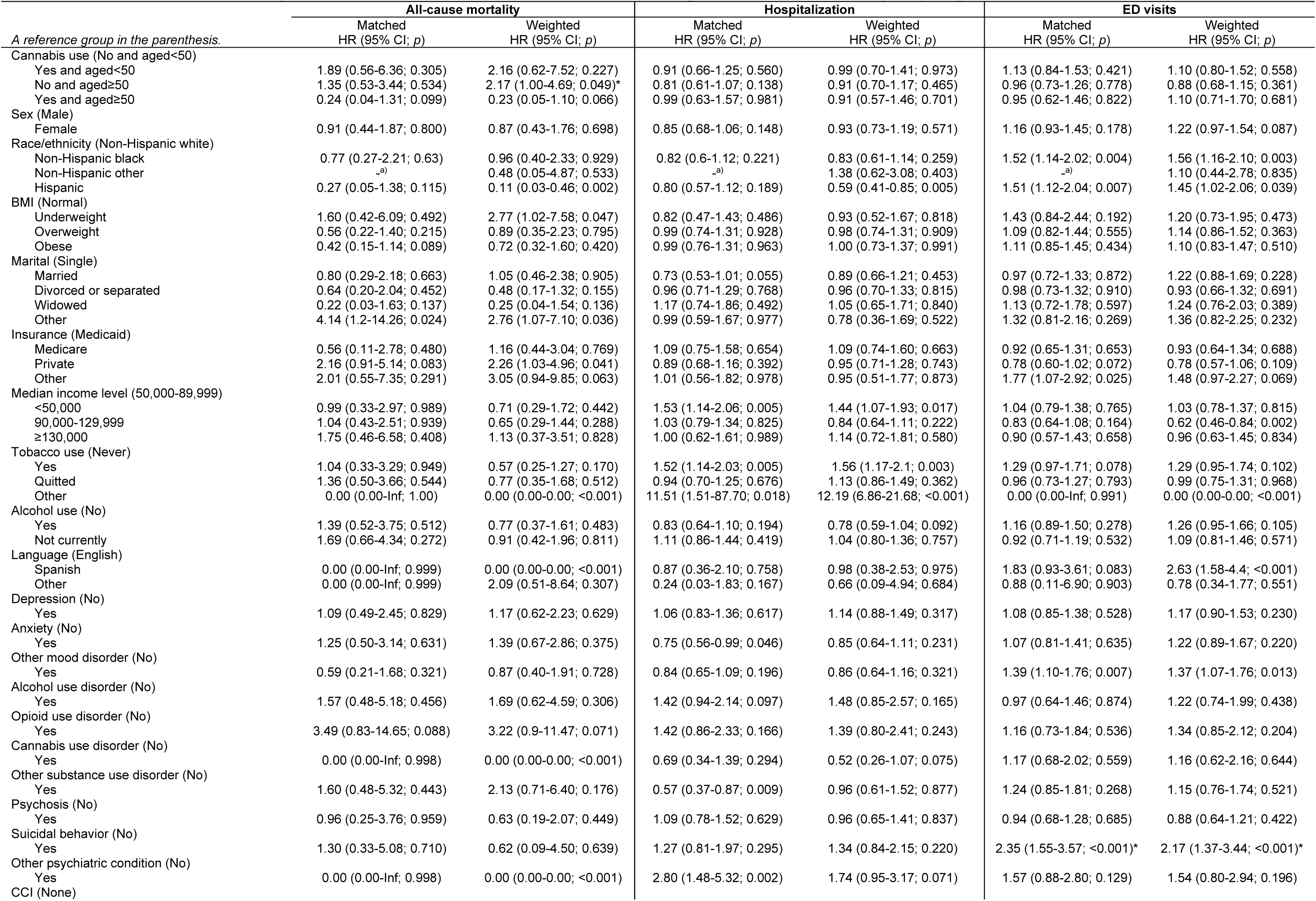
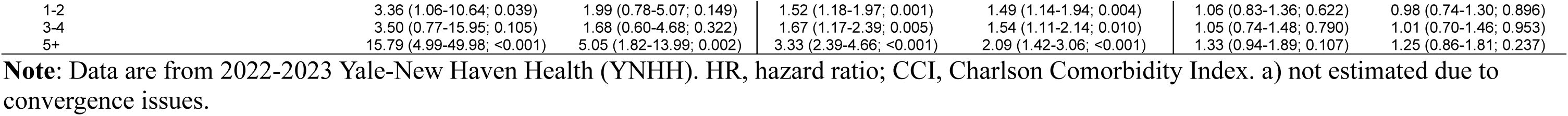
Interaction effects of age and cannabis use on all-cause mortality, hospitalization, and emergency department (ED) visits.

**Table 4.**
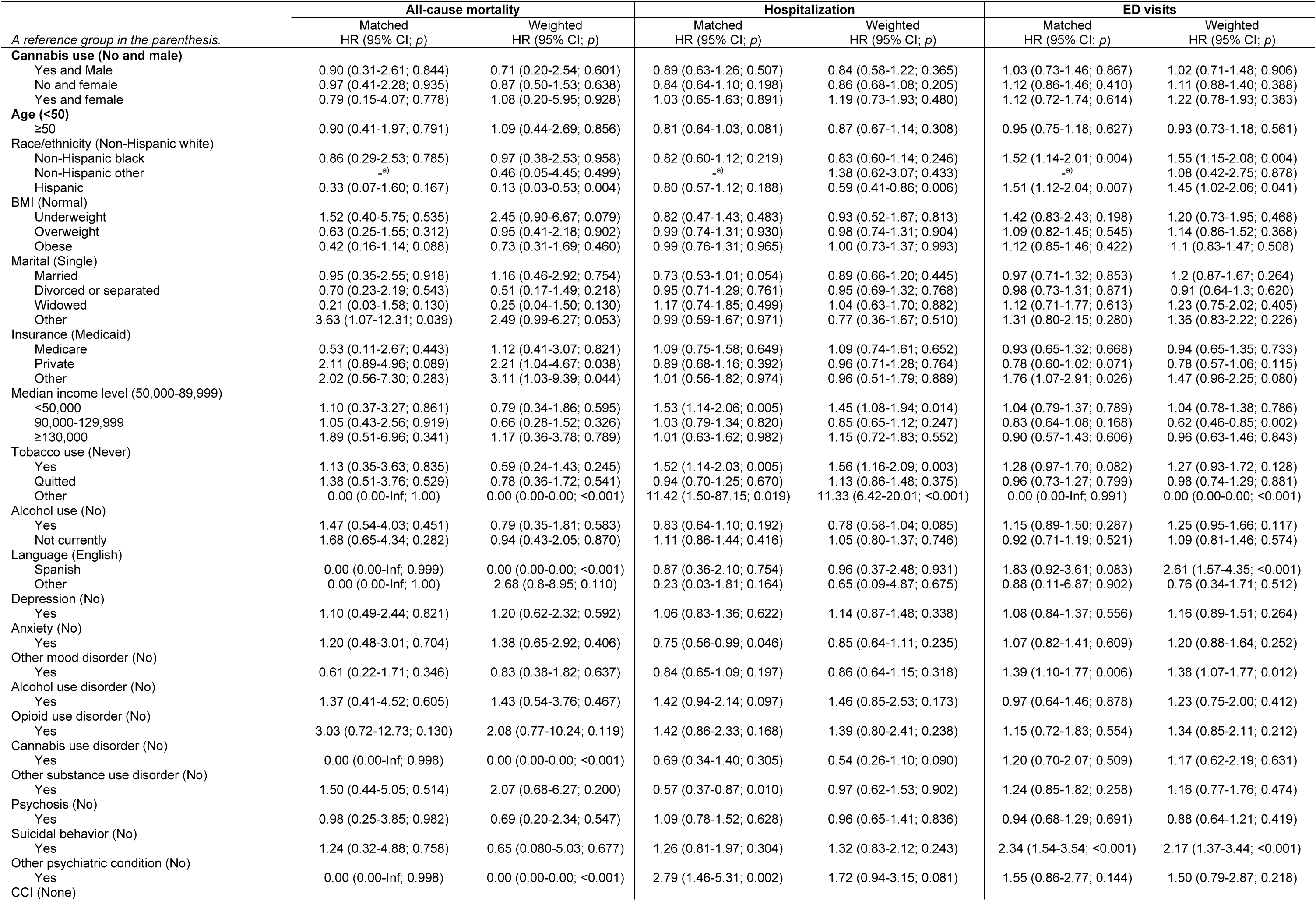
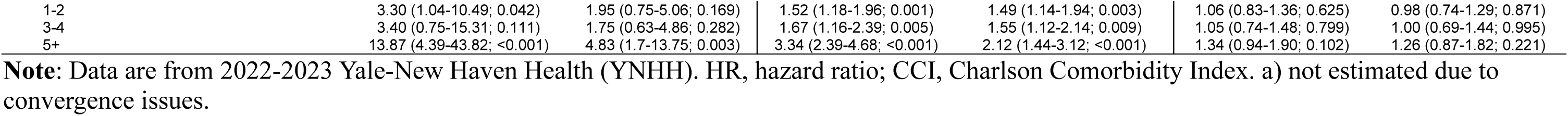
Interaction effects of biological sex and cannabis use on all-cause mortality, hospitalization, and emergency department (ED) visits.

When exploring the impact of cannabis use on change in benzodiazepine dosage over the follow-up period, our analysis revealed that benzodiazepine dosage was overall decreased from 8.13 to 6.65 DME. However, further analysis revealed non-significant group differences.

### Associations of cannabis use with outcomes of interest: Exploratory analyses

In exploratory analysis, we further stratified cannabis use (i.e., medical users, non-medical users, and non-users), we found that compared to non-users, medical cannabis users had a lower risk of all-cause mortality (cHR, 0.08 [0.01-0.88]; *p*=0.039) in the IPTW-adjusted model, but medical cannabis users had a greater risk for hospitalizations (cHR, 1.59 [1.08-2.34]; *p*=0.018) in the PSM-adjusted model (**Table S2**).

In the interaction model of cannabis use and age on the outcomes of interest (**Table S3**), medical cannabis users aged 49 or younger had a greater risk of hospitalization when compared to non-users aged 49 or younger [cHR, 1.79 [1.01-3.17]; *p*=0.047]. Finally, in the interaction model of cannabis use and biological sex on the outcomes of interest (**Table S4**), female medical users had a lower risk of all-cause mortality when compared with male non-users in the IPTW-model; however, we were not able to fit the model given the limited sample size (i.e., no mortality event was observed).

## Discussion

In this study, we examined the impact of cannabis use on health outcomes and benzodiazepine prescriptions among patients receiving benzodiazepine in a longitudinal cohort using electronic health-record (EHR) data at a large healthcare system.

Our results showed that there was no significant association between cannabis use and all-cause mortality, hospitalizations, or emergency department (ED) visits. Cannabis use did not significantly alter prescription benzodiazepine dose. Previous studies have shown a trend toward lower benzodiazepine use with medical cannabis[30, 31], leading to the belief that cannabis has a benzodiazepine-sparing effect. We did not find evidence for this. The discrepant results may be explained by differences in study design [30, 31]. Corroon et al [30]conducted a survey of individuals who were self-selected following convenience sampling (n=1248), and who reported having used cannabis at least once in the previous 90 days. 46% of participants reported using cannabis as a substitute for prescription drugs, including anxiolytics/benzodiazepines (13.6%). Purcell et al, [31]conducted a retrospective analysis in a medical cannabis cohort (n=146). They found that 44.5% of patients had discontinued benzodiazepines at follow-up after 2 medical cannabis prescriptions; and the discontinuation rate remained stable (45.2%) after 3 medical cannabis prescriptions[31]. In contrast to this study, we used a longer period of follow-up of 2 years, complemented by verified pharmacy prescription quantities and confirmation of cannabis status by urine toxicology.

Compared to non-users, medical cannabis use was associated with a significant decrease in all-cause mortality in the unadjusted analysis and after IPTW, though this association was non-significant after PSM. However, younger medical cannabis users (<50 years) demonstrated a significant increase in all-cause mortality compared to young non-cannabis users after matching. Medical cannabis use was associated with an increased risk of hospitalization post-matching, particularly among younger users. However, these results should be interpreted very cautiously due to the small sample size and potential residual confounding.

Regarding ED visits, we found no significant association with cannabis use, either overall or within subgroups stratified by age and sex, suggesting that cannabis use, whether for medical or non-medical purposes, does not appear to substantially influence acute care utilization in this population.

The lack of significant associations between cannabis uses and clinical outcomes in the broader study population may be attributable to several factors. First, the heterogeneity of cannabis formulations, dosages, and routes of administration complicates the assessment of its therapeutic effects [32]. Second, our dataset did not distinguish between the timing, frequency and quantity of use, or purpose of cannabis use (medical vs. recreational), limiting our ability to adjust for these potential confounders [33]. Third, baseline imbalances in demographic and clinical characteristics between cannabis users and non-users, despite adjustment, may have introduced residual confounding [34].

It is important to acknowledge the limitations of the study design in answering the causal questions. Our study utilized a retrospective follow-up study design which has inherent limitations [35]. Even after using matching some of the baseline variables remain unadjusted potentially introducing confounding [34]. Further, we used a single instance of positive cannabis urine toxicology to confirm the use of cannabis. While this can be taken as a proxy, it does not provide information about the longitudinal use patterns of cannabis. Further, this method of classification may have produced errors in categorizing patients into the specified groups (e.g., for those who used cannabis but did not have a urine toxicology reported because it was not clinically indicated at the given encounter). Another weakness is that the index date was chosen based on availability of medical cannabis data rather than a medically relevant event. Additionally, the follow-up period of 2 years in our study may have been insufficient to capture true cohort effects related to cannabis use.

This study contributes to the growing body of literature on the interplay between cannabis use and benzodiazepine-related public health outcomes. While our results do not support that concomitant use cannabis with benzodiazepines improve benzodiazepine associated public health outcomes, this underscores the need for further research to identify patient populations that may derive benefit from cannabis use as an adjunct or alternative to benzodiazepines[36]. Future studies should also aim to differentiate between medical and recreational cannabis use, assess the role of specific cannabinoids (e.g., CBD vs. THC), and explore the long-term effects of cannabis use on mental health and benzodiazepine dependence.

## Data Availability

All data produced in the present study are available upon reasonable request to the authors

## Acknowledgements and disclosures

### Primary Funding

The study was funded by the National Institute of Drug Abuse (R21DA057540).

## Acknowledgments

We acknowledge and appreciate the help of Soundari Sureshanand from Yale Joint Data Analytics Team (JDAT) in data curation.

