## Supplement for "Impact of cannabis use on health outcomes and prescription benzodiazepine use"

### Supplemental Online Content

Singh N, Dai Y, Wilkinson ST, Rhee TG, & Radhakrishnan R. Association of cannabis use with clinical outcomes and pattern of prescription benzodiazepine use. *Addiction*.


**Figure S1.** Flowchart of the cohort selection

**Figure S2**. Kaplan-Meier survival curves for all-cause mortality, hospitalization, and emergency department (ED) visits by cannabis use according to age and biological sex among benzodiazepine users after inverse probability of treatment weighting (IPTW).

**Table S1.** Baseline socio-demographic and clinical characteristics of the study sample by cannabis use status after inverse probability of treatment weighting (IPTW)

**Table S2.** Medical and non-medical cannabis use and its association with all-cause mortality, hospitalization, and emergency department (ED) visits

**Table S3**. Interaction effects of age and medical and non-medical cannabis use on all-cause mortality, hospitalization, and emergency department (ED) visits

**Table S4.** Interaction effects of biological sex and medical and non-medical cannabis use on all-cause mortality, hospitalization, and emergency department (ED) visits

**References**.


This supplemental material has been provided by the authors to give readers additional information about their work.

**Figure S1**. Flowchart of the cohort selection

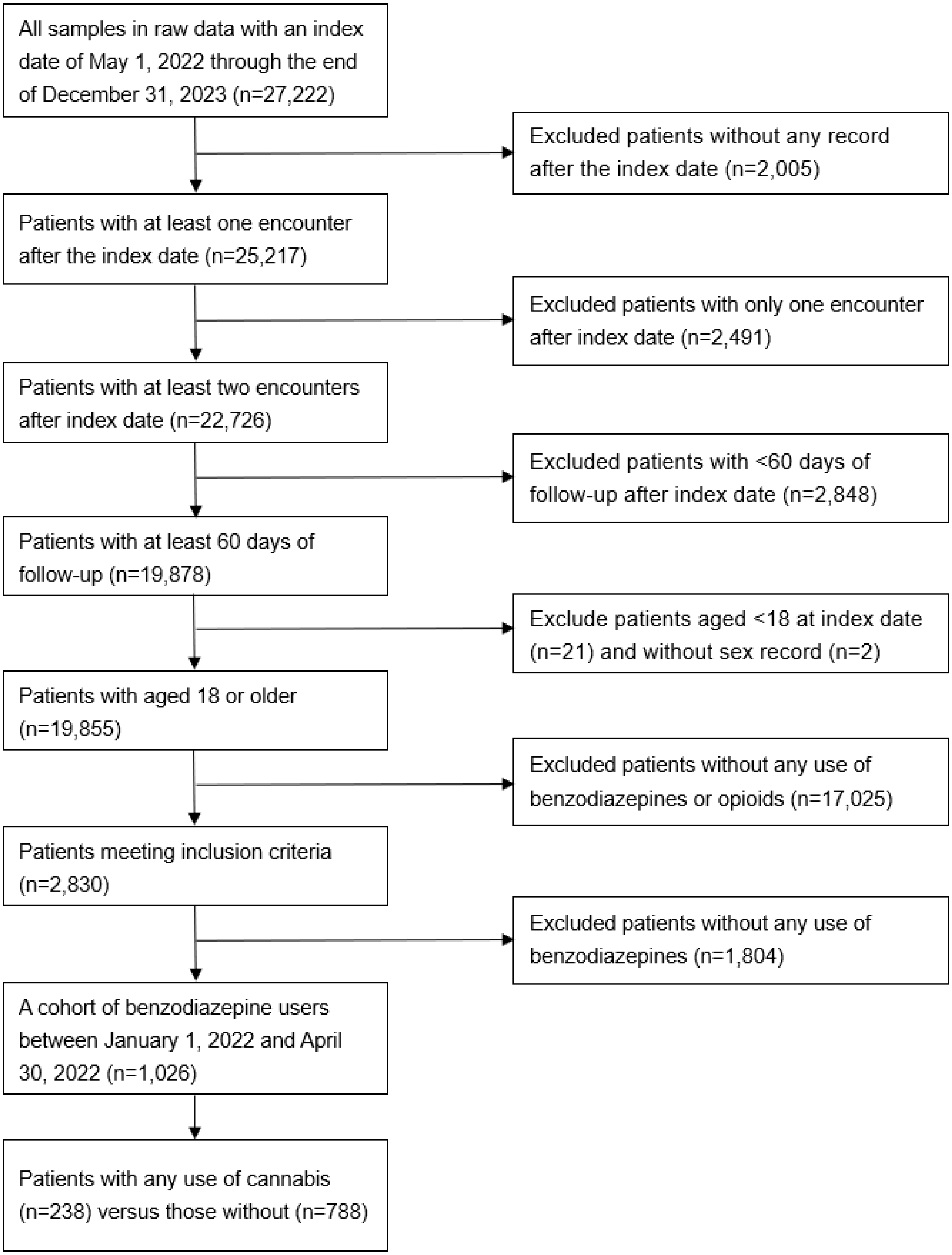


**Note**: Data are from Yale-New Haven Health Systems, 2022-2023.

**Figure S2**. Kaplan-Meier survival curves for all-cause mortality, hospitalization, and emergency department (ED) visits by cannabis use according to age and biological sex among benzodiazepine users after inverse probability of treatment weighting (IPTW).

**Panel 2A**. All-cause mortality by cannabis use and age

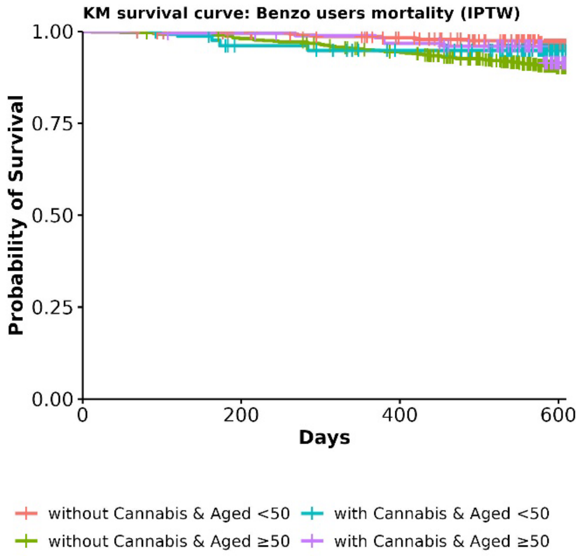


**Panel 2B**. Hospitalization by cannabis use and age

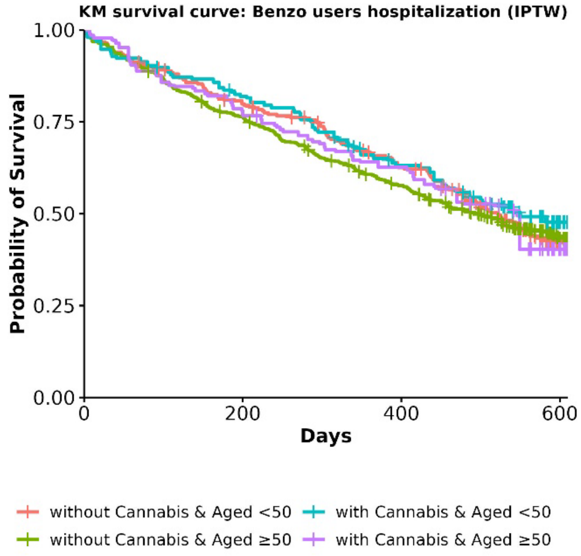


**Panel 2C**. ED visits by cannabis use and age

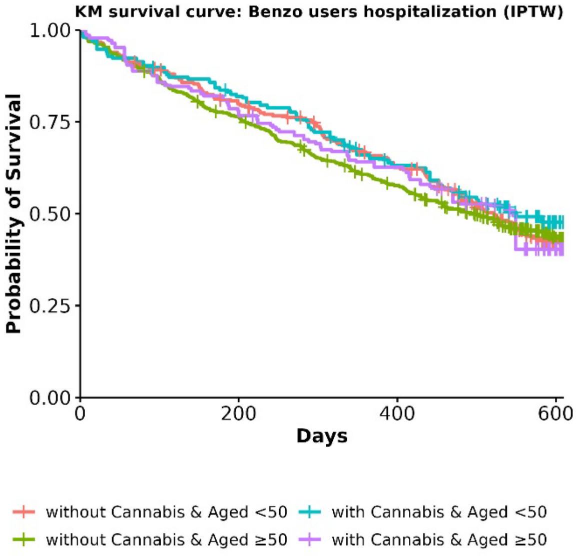


**Panel 2D**. All-cause mortality by cannabis use and biological sex

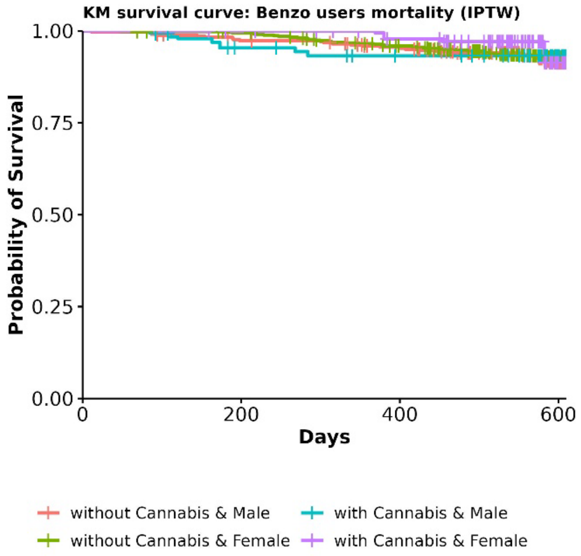


**Panel 2E**. Hospitalization by cannabis use and biological sex

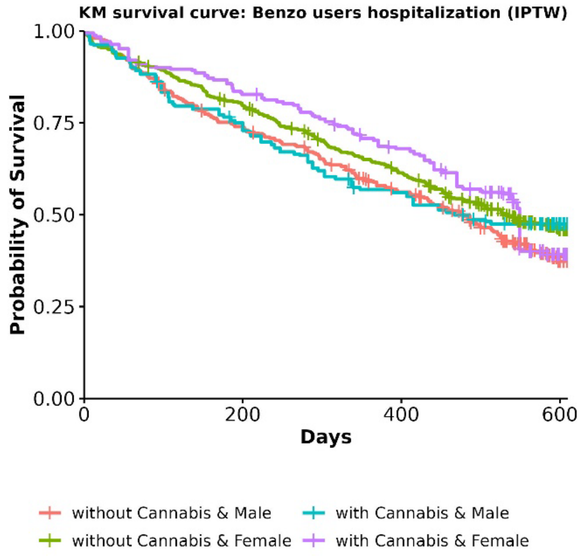


**Panel 2F**. ED visits by cannabis use and age

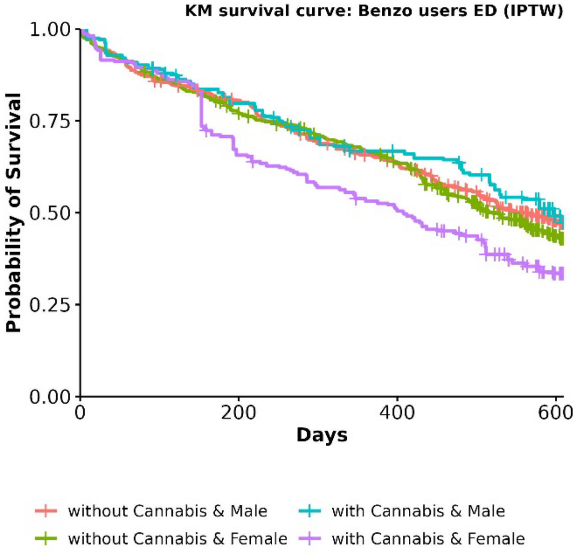

**Table S1.** Baseline socio-demographic and clinical characteristics of the study sample by cannabis use status after inverse probability of treatment weighting (IPTW)

| | After IPTW | | | |
|---|---|---|---|---|
| | **Non-user (n=1,025)** | **User (n=1,016)** | **Total (n=2,041)** | ***P*-value** |
| Age, mean ± standard deviation (SD) | 54.2 ± 16.4 | 53.4 ± 13.8 | 53.8 ± 15.2 | 0.437 |
| Biological sex | | | | |
| Male | 405 (39.5%) | 382 (37.6%) | 787 (38.5%) | 0.365 |
| Female | 620 (60.5%) | 635 (62.4%) | 1255 (61.5%) | |
| Race and ethnicity | | | | |
| Non-Hispanic white | 712 (69.4%) | 680 (66.9%) | 1392 (68.2%) | <0.001 |
| Non-Hispanic black | 133 (13.0%) | 123 (12.1%) | 256 (12.6%) | |
| Hispanic | 144 (14.0%) | 140 (13.7%) | 284 (13.9%) | |
| Non-Hispanic other | 15 (1.5%) | 0 (0.0%) | 15 (0.7%) | |
| Unknown/missing | 21 (2.1%) | 74 (7.2%) | 95 (4.6%) | |
| Marital status | | | | |
| Single | 464 (45.3%) | 463 (45.5%) | 927 (45.4%) | 0.995 |
| Married | 287 (28.0%) | 280 (27.5%) | 566 (27.7%) | |
| Divorced or separated | 163 (15.9%) | 166 (16.4%) | 330 (16.2%) | |
| Widowed | 73 (7.1%) | 72 (7.1%) | 145 (7.1%) | |
| Other | 38 (3.7%) | 35 (3.5%) | 73 (3.6%) | |
| Income level | | | | |
| <50,000 | 160 (15.6%) | 140 (13.7%) | 299 (14.7%) | 0.013 |
| 50,000-89,999 | 426 (41.5%) | 497 (48.9%) | 923 (45.2%) | |
| 90,000-129,999 | 342 (33.3%) | 283 (27.9%) | 625 (30.6%) | |
| 130,000-149,999 | 47 (4.6%) | 50 (5.0%) | 97 (4.8%) | |
| ≥150,000 | 51 (5.0%) | 46 (4.5%) | 97 (4.7%) | |
| Insurance coverage | | | | |
| Private | 383 (37.3%) | 383 (37.7%) | 765 (37.5%) | 0.536 |
| Medicaid | 420 (41.0%) | 390 (38.4%) | 810 (39.7%) | |
| Medicare | 168 (16.3%) | 192 (18.9%) | 360 (17.6%) | |
| Other | 40 (3.9%) | 35 (3.4%) | 74 (3.6%) | |
| Unknown/missing | 15 (1.4%) | 17 (1.6%) | 31 (1.5%) | |
| Primary language use | | | | |
| English | 996 (97.2%) | 998 (98.2%) | 1993 (97.7%) | 0.200 |
| Spanish | 23 (2.2%) | 16 (1.6%) | 39 (1.9%) | |
| Other | 6 (0.6%) | 2 (0.2%) | 9 (0.4%) | |
| BMI, mean ± SD | 29.4 ± 8.0 | 29.5 ± 7.8 | 29.4 ± 7.9 | 0.769 |
| Charlson comorbidity index | | | | |
| None | 477 (46.5%) | 487 (48.0%) | 964 (47.2%) | 0.913 |
| 1-2 | 299 (29.1%) | 293 (28.8%) | 592 (29.0%) | |
| 3-4 | 122 (11.9%) | 116 (11.4%) | 238 (11.7%) | |
| ≥5 | 128 (12.5%) | 120 (11.8%) | 248 (12.1%) | |
| Current alcohol use | | | | |
| No | 389 (37.9%) | 380 (37.4%) | 769 (37.7%) | 0.560 |
| Yes | 313 (30.5%) | 335 (33.0%) | 648 (31.8%) | |
| Not currently | 315 (30.7%) | 295 (29.0%) | 610 (29.9%) | |
| Unknown/missing | 9 (0.9%) | 6 (0.6%) | 15 (0.7%) | |
| Current tobacco use | | | | |
| Never | 385 (37.5%) | 197 (19.4%) | 582 (28.5%) | <0.001 |
| Yes | 265 (25.9%) | 395 (38.8%) | 660 (32.3%) | |
| Quitted | 373 (36.4%) | 424 (41.8%) | 798 (39.1%) | |
| Other | 2 (0.2%) | 0 (0.0%) | 2 (0.1%) | |
| Depression | | | | |
| No | 295 (28.8%) | 265 (26.0%) | 560 (27.4%) | 0.168 |
| Yes | 730 (71.2%) | 752 (74.0%) | 1481 (72.6%) | |
| Anxiety | | | | |
| No | 815 (79.5%) | 773 (76.1%) | 1588 (77.8%) | 0.062 |
| Yes | 210 (20.5%) | 243 (23.9%) | 453 (22.2%) | |
| Other mood disorder | | | | |

| | | | | |
|---|---|---|---|---|
| No | 739 (72.0%) | 733 (72.1%) | 1472 (72.1%) | 0.953 |
| Yes | 287 (28.0%) | 283 (27.9%) | 570 (27.9%) | |
| Suicidal symptoms | | | | |
| No | 964 (94.0%) | 963 (94.8%) | 1927 (94.4%) | 0.470 |
| Yes | 61 (6.0%) | 53 (5.2%) | 114 (5.6%) | |
| Psychosis | | | | |
| No | 904 (88.2%) | 905 (89.0%) | 1809 (88.6%) | 0.531 |
| Yes | 121 (11.8%) | 111 (11.0%) | 232 (11.4%) | |
| Alcohol use disorder | | | | |
| No | 943 (92.0%) | 924 (90.9%) | 1868 (91.5%) | 0.728 |
| Yes | 82 (8.0%) | 92 (9.1%) | 174 (8.5%) | |
| Opioid use disorder | | | | |
| No | 953 (93.0%) | 945 (93.0%) | 1899 (93.0%) | 0.974 |
| Yes | 72 (7.0%) | 71 (7.0%) | 143 (7.0%) | |
| Cannabis use disorder | | | | |
| No | 998 (97.4%) | 985 (96.9%) | 1983 (97.2%) | 0.571 |
| Yes | 27 (2.6%) | 31 (3.1%) | 58 (2.8%) | |
| Other substance use disorder | | | | |
| No | 927 (90.5%) | 905 (89.1%) | 1833 (89.8%) | 0.309 |
| Yes | 98 (9.5%) | 111 (10.9%) | 208 (10.2%) | |
| Other psychiatric condition | | | | |
| No | 991 (96.7%) | 982 (96.6%) | 1973 (96.7%) | 0.971 |
| Yes | 34 (3.3%) | 34 (3.4%) | 68 (3.3%) | |

**Note**: Data are from 2022-2023 Yale-New Haven Health (YNHH). Characteristics before IPTW are reported in Table 1.

**Table S2.** Medical and non-medical cannabis use and its association with all-cause mortality, hospitalization, and emergency department (ED) visits

| | **All-cause mortality** | | **Hospitalization** | | **ED visits** | |
|---|---|---|---|---|---|---|
| | Matched | Weighted | Matched | Weighted | Matched | Weighted |
| *A reference group in the parenthesis.* | HR (95% CI; *p*) | HR (95% CI; *p*) | HR (95% CI; *p*) | HR (95% CI; *p*) | HR (95% CI; *p*) | HR (95% CI; *p*) |
| Cannabis use (No) | | | | | | |
| Yes (Non-medical) | 1.41 (0.62-3.23; 0.416) | 1.13 (0.54-2.37; 0.744) | 0.80 (0.62-1.03; 0.083) | 0.86 (0.65-1.14; 0.291) | 1.08 (0.86-1.37; 0.505) | 1.17 (0.92-1.49; 0.189) |
| Yes (Medical) | 0.14 (0.02-1.05; 0.056) | 0.08 (0.01-0.88; 0.039) | 1.59 (1.08-2.34; 0.018) | 1.31 (0.86-2.00; 0.213) | 1.27 (0.85-1.90; 0.249) | 1.16 (0.75-1.78; 0.506) |
| Age (18-30) | | | | | | |
| 31-49 | 4.01 (0.47-34.14; 0.203) | 5.12 (0.57-46.29; 0.146) | 0.95 (0.66-1.35; 0.762) | 0.86 (0.59-1.26; 0.453) | 0.89 (0.63-1.25; 0.493) | 0.75 (0.51-1.09; 0.128) |
| 50+ | 3.27 (0.37-29.13; 0.288) | 5.20 (0.56-48.01; 0.146) | 0.80 (0.55-1.17; 0.254) | 0.77 (0.53-1.11; 0.162) | 0.82 (0.57-1.18; 0.285) | 0.73 (0.50-1.07; 0.102) |
| Sex (Male) | | | | | | |
| Female | 0.63 (0.3-1.34; 0.229) | 0.72 (0.38-1.34; 0.298) | 0.88 (0.70-1.10; 0.250) | 0.96 (0.76-1.22; 0.742) | 1.08 (0.87-1.35; 0.476) | 1.21 (0.97-1.52; 0.096) |
| Race/ethnicity (Non-Hispanic white) | | | | | | |
| Non-Hispanic black | 0.70 (0.23-2.08; 0.516) | 0.88 (0.34-2.26; 0.784) | 0.89 (0.65-1.21; 0.449) | 0.85 (0.62-1.16; 0.306) | 1.60 (1.20-2.12; 0.001) | 1.56 (1.17-2.10; 0.003) |
| Non-Hispanic other | 0.00 (0.00-Inf; 1.00) | 0.39 (0.04-3.82; 0.421) | 8.41 (1.93-36.66; 0.005) | 1.42 (0.63-3.20; 0.396) | 6.86 (1.59-29.59; 0.010) | 1.08 (0.43-2.73; 0.871) |
| Hispanic | 0.36 (0.07-1.74; 0.201) | 0.10 (0.02-0.40; 0.001) | 0.79 (0.56-1.09; 0.154) | 0.60 (0.42-0.86; 0.006) | 1.54 (1.15-2.07; 0.004) | 1.44 (1.01-2.04; 0.045) |
| BMI (Normal) | | | | | | |
| Underweight | 1.80 (0.49-6.61; 0.374) | 1.58 (0.51-4.87; 0.429) | 0.99 (0.59-1.67; 0.972) | 0.96 (0.54-1.70; 0.886) | 1.27 (0.74-2.16; 0.386) | 1.21 (0.74-1.97; 0.447) |
| Overweight | 0.56 (0.22-1.42; 0.224) | 0.75 (0.33-1.67; 0.474) | 1.01 (0.75-1.34; 0.97) | 1.02 (0.77-1.36; 0.886) | 1.09 (0.82-1.44; 0.572) | 1.15 (0.86-1.54; 0.332) |
| Obese | 0.45 (0.17-1.20; 0.112) | 0.59 (0.27-1.31; 0.193) | 1.00 (0.76-1.32; 0.992) | 1.04 (0.76-1.42; 0.827) | 1.09 (0.83-1.42; 0.532) | 1.11 (0.83-1.49; 0.474) |
| Marital (Single) | | | | | | |
| Married | 0.62 (0.22-1.79; 0.382) | 0.82 (0.35-1.90; 0.636) | 0.86 (0.62-1.20; 0.376) | 0.93 (0.69-1.27; 0.663) | 1.13 (0.83-1.55; 0.435) | 1.26 (0.90-1.77; 0.187) |
| Divorced or separated | 0.71 (0.24-2.13; 0.546) | 0.33 (0.10-1.05; 0.061) | 1.01 (0.75-1.36; 0.939) | 1.00 (0.73-1.38; 0.985) | 1.03 (0.77-1.39; 0.823) | 0.96 (0.68-1.37; 0.830) |
| Widowed | 0.21 (0.03-1.50; 0.120) | 0.27 (0.06-1.29; 0.102) | 1.29 (0.82-2.02; 0.278) | 1.10 (0.68-1.80; 0.694) | 1.27 (0.81-2.00; 0.300) | 1.27 (0.77-2.10; 0.342) |
| Other | 3.55 (1.07-11.78; 0.038) | 2.94 (1.33-6.54; 0.008) | 0.92 (0.52-1.63; 0.782) | 0.71 (0.31-1.66; 0.432) | 1.21 (0.72-2.04; 0.462) | 1.40 (0.84-2.33; 0.193) |
| Insurance (Medicaid) | | | | | | |
| Medicare | 1.01 (0.24-4.16; 0.993) | 1.26 (0.46-3.41; 0.656) | 0.91 (0.63-1.32; 0.624) | 1.08 (0.73-1.58; 0.706) | 0.77 (0.53-1.11; 0.156) | 0.93 (0.64-1.34; 0.688) |
| Private | 2.37 (0.97-5.75; 0.057) | 2.63 (1.27-5.42; 0.009) | 0.78 (0.60-1.02; 0.074) | 0.93 (0.69-1.25; 0.643) | 0.76 (0.58-0.99; 0.041) | 0.78 (0.57-1.06; 0.108) |
| Other | 4.00 (1.07-14.99; 0.039) | 4.18 (1.42-12.3; 0.009) | 0.74 (0.40-1.37; 0.333) | 0.87 (0.46-1.63; 0.663) | 1.52 (0.91-2.53; 0.106) | 1.48 (0.96-2.27; 0.073) |
| Median income level (50,000-89,999) | | | | | | |
| <50,000 | 1.95 (0.69-5.50; 0.207) | 1.15 (0.49-2.74; 0.744) | 1.42 (1.06-1.91; 0.018) | 1.37 (1.02-1.85; 0.038) | 1.03 (0.78-1.36; 0.827) | 1.02 (0.77-1.36; 0.876) |
| 90,000-129,999 | 1.15 (0.44-2.95; 0.778) | 0.66 (0.28-1.55; 0.337) | 1.03 (0.79-1.33; 0.853) | 0.85 (0.65-1.11; 0.230) | 0.80 (0.61-1.04; 0.092) | 0.62 (0.46-0.84; 0.002) |
| ≥130,000 | 2.16 (0.60-7.83; 0.241) | 1.34 (0.45-4.05; 0.601) | 1.24 (0.80-1.93; 0.344) | 1.12 (0.70-1.78; 0.632) | 0.90 (0.58-1.41; 0.660) | 0.94 (0.62-1.44; 0.790) |
| Tobacco use (Never) | | | | | | |
| Yes | 0.90 (0.29-2.76; 0.851) | 0.57 (0.24-1.32; 0.187) | 1.43 (1.07-1.92; 0.017) | 1.6 (1.18-2.16; 0.002) | 1.24 (0.93-1.65; 0.139) | 1.34 (0.98-1.81; 0.064) |
| Quitted | 0.91 (0.35-2.41; 0.854) | 0.76 (0.36-1.63; 0.485) | 0.85 (0.63-1.14; 0.269) | 1.13 (0.85-1.49; 0.393) | 0.98 (0.75-1.3; 0.911) | 1.02 (0.77-1.36; 0.868) |
| Other | 0.00 (0.00-Inf; 1.00) | 0.00 (0.00-0.00; <0.001) | 11.11 (1.45-85.06; 0.020) | 12.58 (6.68-23.70; <0.001) | 0.00 (0.00-Inf; 0.991) | 0.00 (0.00-0.00; <0.001) |
| Alcohol use (No) | | | | | | |
| Yes | 1.10 (0.40-2.98; 0.857) | 0.78 (0.37-1.63; 0.504) | 0.86 (0.66-1.13; 0.273) | 0.79 (0.59-1.05; 0.100) | 1.22 (0.94-1.59; 0.133) | 1.26 (0.95-1.66; 0.105) |
| Not currently | 1.55 (0.63-3.81; 0.344) | 0.97 (0.46-2.07; 0.944) | 1.09 (0.84-1.42; 0.504) | 1.05 (0.81-1.38; 0.700) | 1.06 (0.81-1.37; 0.681) | 1.08 (0.80-1.45; 0.617) |
| Language (English) | | | | | | |
| Spanish | 0.00 (0.00-Inf; 0.999) | 0.00 (0.00-0.00; <0.001) | 1.15 (0.49-2.68; 0.747) | 1.02 (0.40-2.64; 0.960) | 1.88 (0.97-3.64; 0.061) | 2.69 (1.62-4.45; <0.001) |
| Other | 0.00 (0.00-Inf; 1.00) | 4.26 (1.19-15.29; 0.026) | 0.15 (0.02-1.19; 0.073) | 0.52 (0.06-4.27; 0.546) | 0.76 (0.09-6.07; 0.794) | 0.81 (0.35-1.86; 0.620) |
| Depression (No) | | | | | | |
| Yes | 1.19 (0.52-2.69; 0.682) | 1.25 (0.65-2.42; 0.501) | 1.10 (0.85-1.40; 0.472) | 1.13 (0.87-1.47; 0.370) | 1.13 (0.89-1.45; 0.314) | 1.17 (0.90-1.52; 0.234) |
| Anxiety (No) | | | | | | |
| Yes | 1.14 (0.46-2.84; 0.781) | 1.34 (0.61-2.90; 0.464) | 0.71 (0.54-0.95; 0.021) | 0.85 (0.64-1.12; 0.248) | 1.12 (0.85-1.47; 0.420) | 1.22 (0.90-1.67; 0.206) |
| Other mood disorder (No) | | | | | | |
| Yes | 0.44 (0.16-1.23; 0.117) | 0.73 (0.33-1.61; 0.432) | 0.91 (0.71-1.18; 0.480) | 0.88 (0.66-1.18; 0.387) | 1.36 (1.07-1.72; 0.012) | 1.37 (1.07-1.75; 0.013) |
| Alcohol use disorder (No) | | | | | | |
| Yes | 1.49 (0.46-4.83; 0.502) | 1.09 (0.41-2.86; 0.866) | 1.36 (0.91-2.04; 0.138) | 1.50 (0.86-2.61; 0.149) | 1.01 (0.68-1.50; 0.960) | 1.23 (0.76-2.01; 0.401) |
| Opioid use disorder (No) | | | | | | |
| Yes | 5.47 (1.21-24.68; 0.027) | 3.11 (0.90-10.72; 0.073) | 1.40 (0.84-2.31; 0.193) | 1.34 (0.76-2.34; 0.311) | 0.95 (0.59-1.51; 0.814) | 1.34 (0.85-2.10; 0.206) |
| Cannabis use disorder (No) | | | | | | |
| Yes | 0.00 (0.00-Inf; 0.998) | 0.00 (0.00-0.00; <0.001) | 0.76 (0.39-1.47; 0.410) | 0.54 (0.27-1.11; 0.093) | 1.24 (0.73-2.10; 0.422) | 1.12 (0.60-2.10; 0.716) |
| Other substance use disorder (No) | | | | | | |
| Yes | 0.99 (0.29-3.42; 0.984) | 2.52 (0.84-7.59; 0.099) | 0.65 (0.43-0.98; 0.042) | 0.93 (0.59-1.48; 0.760) | 1.38 (0.95-2.01; 0.089) | 1.15 (0.76-1.75; 0.501) |
| Psychosis (No) | | | | | | |
| Yes | 1.51 (0.40-5.72; 0.545) | 0.92 (0.31-2.74; 0.876) | 1.03 (0.74-1.45; 0.842) | 0.95 (0.64-1.39; 0.786) | 0.93 (0.68-1.27; 0.645) | 0.88 (0.64-1.21; 0.443) |
| Suicidal behavior (No) | | | | | | |
| Yes | 2.72 (0.73-10.12; 0.135) | 1.22 (0.20-7.44; 0.831) | 1.27 (0.82-1.95; 0.280) | 1.32 (0.83-2.11; 0.241) | 1.87 (1.24-2.84; 0.003) | 2.18 (1.38-3.46; <0.001) |
| Other psychiatric condition (No) | | | | | | |

| | | | | | | |
|---|---|---|---|---|---|---|
| Yes | 0.00 (0.00-Inf; 0.998) | 0.00 (0.00-0.00; <0.001) | 1.93 (1.08-3.45; 0.026) | 1.73 (0.94-3.19; 0.081) | 1.29 (0.75-2.24; 0.357) | 1.54 (0.81-2.94; 0.187) |
| CCI (None) | | | | | | |
| 1-2 | 2.45 (0.80-7.49; 0.117) | 2.09 (0.78-5.59; 0.141) | 1.47 (1.13-1.89; 0.004) | 1.47 (1.12-1.92; 0.005) | 1.01 (0.78-1.29; 0.954) | 0.98 (0.74-1.30; 0.884) |
| 3-4 | 3.31 (0.86-12.83; 0.083) | 1.70 (0.59-4.86; 0.324) | 1.61 (1.13-2.30; 0.008) | 1.55 (1.13-2.14; 0.007) | 1.04 (0.74-1.47; 0.804) | 1.03 (0.71-1.49; 0.895) |
| 5+ | 9.89 (3.30-29.61; <0.001) | 5.52 (1.98-15.33; 0.001) | 2.97 (2.11-4.17; <0.001) | 2.00 (1.36-2.94; <0.001) | 1.42 (1.00-2.01; 0.048) | 1.25 (0.87-1.81; 0.223) |

**Note**: Data are from 2022-2023 Yale-New Haven Health (YNHH). HR, hazard ratio; CCI, Charlson Comorbidity Index. a) not estimated due to convergence issues.

**Table S3**. Interaction effects of age and medical and non-medical cannabis use on all-cause mortality, hospitalization, and emergency department (ED) visits

| | All-cause mortality | | Hospitalization | | ED visits | |
|---|---|---|---|---|---|---|
| | Matched | Weighted | Matched | Weighted | Matched | Weighted |
| *A reference group in the parenthesis.* | HR (95% CI; *p*) | HR (95% CI; *p*) | HR (95% CI; *p*) | HR (95% CI; *p*) | HR (95% CI; *p*) | HR (95% CI; *p*) |
| Cannabis use (No and aged<50) | | | | | | |
| Yes (non-medical) and aged<50 | 2.03 (0.56-7.30; 0.280) | 2.35 (0.66-8.39; 0.189) | 0.82 (0.57-1.16; 0.258) | 0.93 (0.63-1.37; 0.715) | 1.09 (0.79-1.51; 0.582) | 1.11 (0.78-1.57; 0.575) |
| Yes (medical) and aged<50 | 2.48 (0.25-24.08; 0.434) | 1.00 (0.05-20.5; 0.999) | 1.79 (1.01-3.17; 0.047) | 1.44 (0.78-2.67; 0.246) | 1.28 (0.74-2.24; 0.376) | 1.08 (0.61-1.92; 0.782) |
| No and aged≥50 | 1.15 (0.45-2.94; 0.778) | 2.00 (0.94-4.24; 0.071) | 0.86 (0.65-1.14; 0.294) | 0.92 (0.71-1.18; 0.510) | 0.91 (0.70-1.20; 0.513) | 0.88 (0.68-1.15; 0.360) |
| Yes (non-medical) and aged≥50 | 0.62 (0.11-3.50; 0.584) | 0.37 (0.08-1.83; 0.224) | 0.96 (0.58-1.60; 0.888) | 0.88 (0.51-1.51; 0.646) | 0.98 (0.62-1.56; 0.931) | 1.10 (0.68-1.77; 0.709) |
| Yes (medical) and aged≥50 | 0.01 (0.00-0.58; 0.026) | 0.04 (0.00-2.78; 0.136) | 0.82 (0.38-1.76; 0.605) | 0.87 (0.39-1.94; 0.736) | 0.98 (0.44-2.16; 0.953) | 1.10 (0.49-2.50; 0.812) |
| Sex (Male) | | | | | | |
| Female | 0.60 (0.28-1.26; 0.177) | 0.69 (0.37-1.26; 0.225) | 0.87 (0.69-1.09; 0.233) | 0.96 (0.76-1.22; 0.743) | 1.08 (0.87-1.35; 0.472) | 1.22 (0.97-1.53; 0.087) |
| Race/ethnicity (Non-Hispanic white) | | | | | | |
| Non-Hispanic black | 0.63 (0.21-1.88; 0.405) | 0.86 (0.33-2.23; 0.753) | 0.89 (0.65-1.21; 0.442) | 0.85 (0.62-1.16; 0.297) | 1.60 (1.20-2.12; 0.001) | 1.56 (1.16-2.10; 0.003) |
| Non-Hispanic other | 0.00 (0.00-Inf; 1.00) | 0.34 (0.03-3.75; 0.377) | 8.89 (2.07-38.2; 0.003) | 1.46 (0.65-3.28; 0.361) | 7.44 (1.75-31.58; 0.007) | 1.10 (0.44-2.77; 0.838) |
| Hispanic | 0.33 (0.07-1.64; 0.177) | 0.09 (0.02-0.37; <0.001) | 0.78 (0.56-1.09; 0.142) | 0.59 (0.41-0.86; 0.006) | 1.55 (1.15-2.08; 0.004) | 1.45 (1.02-2.07; 0.040) |
| BMI (Normal) | | | | | | |
| Underweight | 1.93 (0.53-7.04; 0.319) | 1.80 (0.57-5.72; 0.318) | 0.98 (0.58-1.66; 0.952) | 0.96 (0.55-1.70; 0.896) | 1.26 (0.74-2.15; 0.401) | 1.20 (0.73-1.95; 0.476) |
| Overweight | 0.56 (0.22-1.39; 0.209) | 0.77 (0.34-1.74; 0.529) | 1.01 (0.75-1.34; 0.972) | 1.01 (0.76-1.35; 0.921) | 1.08 (0.81-1.43; 0.590) | 1.14 (0.85-1.52; 0.370) |
| Obese | 0.49 (0.19-1.31; 0.157) | 0.64 (0.29-1.40; 0.262) | 1.00 (0.76-1.31; 0.997) | 1.03 (0.75-1.41; 0.862) | 1.08 (0.83-1.41; 0.568) | 1.10 (0.82-1.47; 0.520) |
| Marital (Single) | | | | | | |
| Married | 0.65 (0.22-1.93; 0.443) | 0.86 (0.36-2.05; 0.734) | 0.85 (0.61-1.18; 0.341) | 0.92 (0.68-1.24; 0.582) | 1.11 (0.82-1.51; 0.512) | 1.22 (0.87-1.70; 0.243) |
| Divorced or separated | 0.72 (0.24-2.16; 0.553) | 0.33 (0.10-1.10; 0.071) | 1.00 (0.75-1.34; 0.986) | 0.99 (0.72-1.37; 0.972) | 1.02 (0.76-1.36; 0.917) | 0.93 (0.66-1.32; 0.688) |
| Widowed | 0.34 (0.05-2.21; 0.256) | 0.33 (0.06-1.67; 0.179) | 1.28 (0.81-2.00; 0.291) | 1.10 (0.67-1.79; 0.709) | 1.25 (0.80-1.96; 0.333) | 1.24 (0.75-2.04; 0.399) |
| Other | 4.22 (1.25-14.24; 0.021) | 3.33 (1.38-8.06; 0.007) | 0.93 (0.52-1.64; 0.797) | 0.71 (0.31-1.65; 0.431) | 1.20 (0.72-2.01; 0.492) | 1.36 (0.82-2.25; 0.233) |
| Insurance (Medicaid) | | | | | | |
| Medicare | 1.04 (0.25-4.35; 0.954) | 1.27 (0.48-3.34; 0.635) | 0.91 (0.63-1.33; 0.641) | 1.08 (0.74-1.59; 0.679) | 0.76 (0.53-1.10; 0.152) | 0.93 (0.64-1.34; 0.692) |
| Private | 2.53 (1.06-6.03; 0.036) | 2.59 (1.26-5.32; 0.009) | 0.79 (0.60-1.03; 0.086) | 0.93 (0.69-1.26; 0.658) | 0.76 (0.58-0.99; 0.043) | 0.78 (0.57-1.06; 0.113) |
| Other | 4.05 (1.07-15.39; 0.040) | 3.83 (1.23-11.94; 0.021) | 0.75 (0.40-1.39; 0.356) | 0.86 (0.46-1.62; 0.652) | 1.52 (0.91-2.53; 0.109) | 1.49 (0.96-2.30; 0.072) |
| Median income level (50,000-89,999) | | | | | | |
| <50,000 | 1.95 (0.68-5.56; 0.212) | 1.06 (0.46-2.41; 0.895) | 1.43 (1.07-1.92; 0.017) | 1.38 (1.02-1.86; 0.036) | 1.04 (0.79-1.36; 0.796) | 1.04 (0.78-1.38; 0.813) |
| 90,000-129,999 | 1.07 (0.41-2.77; 0.892) | 0.61 (0.26-1.43; 0.259) | 1.02 (0.79-1.33; 0.862) | 0.84 (0.64-1.10; 0.214) | 0.80 (0.61-1.04; 0.095) | 0.62 (0.46-0.84; 0.002) |
| ≥130,000 | 2.03 (0.54-7.64; 0.297) | 1.26 (0.43-3.73; 0.674) | 1.25 (0.80-1.94; 0.331) | 1.11 (0.70-1.77; 0.647) | 0.91 (0.59-1.42; 0.688) | 0.96 (0.63-1.46; 0.838) |
| Tobacco use (Never) | | | | | | |
| Yes | 1.01 (0.33-3.06; 0.985) | 0.63 (0.29-1.36; 0.238) | 1.42 (1.06-1.89; 0.017) | 1.56 (1.17-2.10; 0.003) | 1.21 (0.92-1.61; 0.174) | 1.29 (0.95-1.74; 0.102) |
| Quitted | 0.98 (0.37-2.57; 0.966) | 0.83 (0.39-1.77; 0.628) | 0.84 (0.63-1.11; 0.221) | 1.10 (0.84-1.45; 0.478) | 0.96 (0.73-1.27; 0.79) | 1.00 (0.75-1.31; 0.972) |
| Other | 0.00 (0.00-Inf; 1.00) | 0.00 (0.00-0.00; <0.001) | 10.97 (1.44-83.48; 0.021) | 12.17 (6.87-21.56; <0.001) | 0.00 (0.00-Inf; 0.991) | 0.00 (0.00-0.00; <0.001) |
| Alcohol use (No) | | | | | | |
| Yes | 0.94 (0.34-2.57; 0.900) | 0.73 (0.34-1.53; 0.402) | 0.85 (0.65-1.12; 0.260) | 0.79 (0.59-1.05; 0.100) | 1.23 (0.94-1.60; 0.131) | 1.26 (0.95-1.66; 0.103) |
| Not currently | 1.52 (0.61-3.79; 0.307) | 0.96 (0.45-2.06; 0.921) | 1.09 (0.84-1.42; 0.503) | 1.06 (0.81-1.38; 0.685) | 1.06 (0.82-1.38; 0.656) | 1.09 (0.81-1.46; 0.578) |
| Language (English) | | | | | | |
| Spanish | 0.00 (0.00-Inf; 0.999) | 0.00 (0.00-0.00; <0.001) | 1.15 (0.49-2.71; 0.742) | 1.03 (0.40-2.63; 0.949) | 1.87 (0.96-3.61; 0.064) | 2.63 (1.58-4.4; <0.001) |
| Other | 0.00 (0.00-Inf; 1.000) | 3.30 (0.76-14.42; 0.113) | 0.13 (0.02-1.10; 0.062) | 0.48 (0.05-4.35; 0.516) | 0.75 (0.09-6.14; 0.789) | 0.78 (0.33-1.83; 0.574) |
| Depression (No) | | | | | | |
| Yes | 1.12 (0.48-2.58; 0.796) | 1.20 (0.61-2.37; 0.595) | 1.09 (0.85-1.4; 0.496) | 1.13 (0.87-1.48; 0.357) | 1.13 (0.89-1.45; 0.320) | 1.18 (0.90-1.53; 0.232) |
| Anxiety (No) | | | | | | |
| Yes | 1.18 (0.48-2.87; 0.720) | 1.34 (0.63-2.86; 0.445) | 0.71 (0.53-0.94; 0.018) | 0.84 (0.64-1.10; 0.210) | 1.11 (0.84-1.46; 0.456) | 1.22 (0.89-1.67; 0.221) |
| Other mood disorder (No) | | | | | | |
| Yes | 0.39 (0.13-1.14; 0.087) | 0.71 (0.31-1.64; 0.418) | 0.91 (0.71-1.18; 0.488) | 0.89 (0.66-1.18; 0.410) | 1.35 (1.06-1.72; 0.013) | 1.37 (1.06-1.77; 0.016) |
| Alcohol use disorder (No) | | | | | | |
| Yes | 1.82 (0.55-6.01; 0.325) | 1.26 (0.42-3.78; 0.685) | 1.37 (0.91-2.05; 0.131) | 1.51 (0.87-2.61; 0.143) | 1.01 (0.68-1.50; 0.977) | 1.21 (0.74-1.98; 0.439) |
| Opioid use disorder (No) | | | | | | |
| Yes | 6.41 (1.46-28.21; 0.014) | 3.35 (0.96-11.72; 0.058) | 1.40 (0.85-2.32; 0.191) | 1.34 (0.76-2.36; 0.310) | 0.95 (0.60-1.52; 0.836) | 1.35 (0.85-2.12; 0.202) |
| Cannabis use disorder (No) | | | | | | |
| Yes | 0.00 (0.00-Inf; 0.998) | 0.00 (0.00-0.00; <0.001) | 0.74 (0.38-1.45; 0.379) | 0.53 (0.26-1.09; 0.085) | 1.25 (0.74-2.13; 0.403) | 1.16 (0.62-2.16; 0.643) |
| Other substance use disorder (No) | | | | | | |
| Yes | 0.92 (0.27-3.11; 0.893) | 2.38 (0.81-7.02; 0.117) | 0.65 (0.43-0.98; 0.040) | 0.93 (0.58-1.47; 0.745) | 1.38 (0.95-2.01; 0.093) | 1.15 (0.75-1.75; 0.522) |
| Psychosis (No) | | | | | | |
| Yes | 1.43 (0.37-5.60; 0.604) | 0.88 (0.29-2.68; 0.820) | 1.04 (0.74-1.46; 0.836) | 0.95 (0.65-1.40; 0.808) | 0.93 (0.67-1.27; 0.633) | 0.88 (0.64-1.21; 0.422) |
| Suicidal behavior (No) | | | | | | |
| Yes | 3.32 (0.91-12.05; 0.068) | 1.39 (0.23-8.46; 0.722) | 1.26 (0.82-1.94; 0.285) | 1.33 (0.83-2.11; 0.237) | 1.87 (1.23-2.83; 0.003) | 2.17 (1.37-3.44; <0.001) |

| | | | | | | |
|---|---|---|---|---|---|---|
| Other psychiatric condition (No) | | | | | | |
| Yes | 0.00 (0.00-Inf; 0.998) | 0.00 (0.00-0.00; <0.001) | 1.95 (1.09-3.49; 0.024) | 1.75 (0.95-3.21; 0.073) | 1.29 (0.74-2.23; 0.370) | 1.53 (0.80-2.94; 0.196) |
| CCI (None) | | | | | | |
| 1-2 | 2.58 (0.83-7.99; 0.100) | 2.02 (0.75-5.43; 0.164) | 1.46 (1.13-1.89; 0.004) | 1.45 (1.11-1.91; 0.007) | 1.01 (0.78-1.29; 0.963) | 0.98 (0.74-1.3; 0.900) |
| 3-4 | 3.89 (0.98-15.46; 0.054) | 1.74 (0.6-5.09; 0.311) | 1.61 (1.13-2.28; 0.008) | 1.52 (1.11-2.10; 0.010) | 1.04 (0.74-1.46; 0.832) | 1.01 (0.7-1.47; 0.949) |
| 5+ | 12.17 (3.96-37.44; <0.001) | 5.91 (2.14-16.34; <0.001) | 3.00 (2.13-4.22; <0.001) | 1.99 (1.35-2.94; <0.001) | 1.42 (1.00-2.01; 0.050) | 1.25 (0.87-1.8; 0.229) |

**Note**: Data are from 2022-2023 Yale-New Haven Health (YNHH). HR, hazard ratio; CCI, Charlson Comorbidity Index. a) not estimated due to convergence issues.

**Table S4.** Interaction effects of biological sex and medical and non-medical cannabis use on all-cause mortality, hospitalization, and emergency department (ED) visits

| | All-cause mortality | | Hospitalization | | ED visits | |
|---|---|---|---|---|---|---|
| | Matched HR (95% CI; *p*) | Weighted HR (95% CI; *p*) | Matched HR (95% CI; *p*) | Weighted HR (95% CI; *p*) | Matched HR (95% CI; *p*) | Weighted HR (95% CI; *p*) |
| Cannabis use (No and male) | | | | | | |
| Yes (non-medical) and male | 1.89 (0.60-6.01; 0.278) | 1.46 (0.45-4.66; 0.527) | 0.74 (0.49-1.12; 0.153) | 0.75 (0.48-1.16; 0.189) | 0.92 (0.62-1.36; 0.673) | 0.99 (0.65-1.51; 0.967) |
| Yes (medical) and male | 0.18 (0.02-1.47; 0.109) | 0.10 (0.01-1.41; 0.088) | 1.51 (0.90-2.53; 0.114) | 1.11 (0.60-2.07; 0.741) | 1.17 (0.68-2.03; 0.566) | 1.09 (0.59-2.01; 0.777) |
| No and female | 0.77 (0.32-1.83; 0.553) | 0.83 (0.46-1.49; 0.533) | 0.84 (0.64-1.10; 0.213) | 0.86 (0.68-1.08; 0.196) | 1.00 (0.77-1.30; 0.983) | 1.11 (0.88-1.40; 0.393) |
| Yes (non-medical) and female | 0.58 (0.11-3.10; 0.524) | 0.67 (0.15-3.01; 0.602) | 1.13 (0.67-1.90; 0.639) | 1.25 (0.72-2.17; 0.418) | 1.29 (0.80-2.09; 0.301) | 1.27 (0.76-2.13; 0.358) |
| Yes (medical) and female | 0.00 (0.00-Inf; 0.999) | 0.00 (0.00-0.00; <0.001) | 1.11 (0.51-2.41; 0.785) | 1.44 (0.67-3.09; 0.355) | 1.16 (0.52-2.59; 0.726) | 1.08 (0.48-2.41; 0.849) |
| Age (<50) | | | | | | |
| ≥50 | 0.92 (0.41-2.06; 0.839) | 1.21 (0.53-2.76; 0.657) | 0.84 (0.66-1.07; 0.158) | 0.87 (0.66-1.13; 0.299) | 0.91 (0.73-1.14; 0.407) | 0.93 (0.73-1.18; 0.539) |
| Race/ethnicity (Non-Hispanic white) | | | | | | |
| Non-Hispanic black | 0.76 (0.25-2.29; 0.627) | 0.92 (0.35-2.42; 0.866) | 0.88 (0.65-1.21; 0.436) | 0.84 (0.61-1.15; 0.268) | 1.59 (1.19-2.11; 0.002) | 1.55 (1.15-2.08; 0.004) |
| Non-Hispanic other | 0.00 (0.00-Inf; 1) | 0.34 (0.04-3.32; 0.356) | 8.85 (2.06-37.98; 0.003) | 1.45 (0.65-3.24; 0.366) | 7.59 (1.79-32.15; 0.006) | 1.08 (0.43-2.75; 0.871) |
| Hispanic | 0.36 (0.07-1.77; 0.208) | 0.10 (0.03-0.41; 0.001) | 0.79 (0.56-1.10; 0.155) | 0.59 (0.41-0.86; 0.006) | 1.55 (1.15-2.09; 0.004) | 1.45 (1.02-2.06; 0.040) |
| BMI (Normal) | | | | | | |
| Underweight | 1.96 (0.55-6.97; 0.297) | 1.63 (0.53-5.02; 0.391) | 0.99 (0.58-1.67; 0.957) | 0.96 (0.54-1.69; 0.879) | 1.26 (0.74-2.15; 0.397) | 1.21 (0.74-1.96; 0.449) |
| Overweight | 0.60 (0.24-1.50; 0.275) | 0.81 (0.38-1.76; 0.595) | 1.01 (0.75-1.34; 0.969) | 1.02 (0.76-1.35; 0.915) | 1.09 (0.82-1.45; 0.548) | 1.14 (0.85-1.53; 0.367) |
| Obese | 0.49 (0.18-1.29; 0.146) | 0.65 (0.29-1.46; 0.297) | 1.00 (0.76-1.31; 0.998) | 1.03 (0.76-1.41; 0.849) | 1.09 (0.84-1.42; 0.522) | 1.10 (0.82-1.48; 0.511) |
| Marital (Single) | | | | | | |
| Married | 0.73 (0.25-2.12; 0.567) | 0.90 (0.36-2.27; 0.822) | 0.85 (0.61-1.18; 0.341) | 0.92 (0.68-1.25; 0.611) | 1.10 (0.81-1.50; 0.541) | 1.20 (0.86-1.68; 0.284) |
| Divorced or separated | 0.78 (0.26-2.34; 0.662) | 0.35 (0.11-1.16; 0.087) | 1.00 (0.75-1.34; 0.991) | 0.99 (0.71-1.37; 0.938) | 1.00 (0.74-1.33; 0.974) | 0.91 (0.64-1.30; 0.612) |
| Widowed | 0.22 (0.03-1.61; 0.137) | 0.29 (0.06-1.49; 0.138) | 1.27 (0.81-2.00; 0.300) | 1.09 (0.66-1.81; 0.728) | 1.24 (0.79-1.94; 0.350) | 1.24 (0.75-2.03; 0.405) |
| Other | 3.79 (1.14-12.61; 0.03) | 3.03 (1.25-7.36; 0.014) | 0.91 (0.52-1.62; 0.759) | 0.72 (0.32-1.63; 0.431) | 1.19 (0.71-1.99; 0.518) | 1.34 (0.82-2.21; 0.246) |
| Insurance (Medicaid) | | | | | | |
| Medicare | 0.94 (0.23-3.92; 0.937) | 1.18 (0.43-3.24; 0.742) | 0.91 (0.63-1.33; 0.639) | 1.09 (0.74-1.60; 0.657) | 0.77 (0.53-1.12; 0.167) | 0.93 (0.65-1.35; 0.717) |
| Private | 2.23 (0.94-5.30; 0.069) | 2.56 (1.27-5.18; 0.009) | 0.79 (0.60-1.03; 0.080) | 0.94 (0.70-1.27; 0.688) | 0.76 (0.58-0.99; 0.041) | 0.78 (0.57-1.06; 0.109) |
| Other | 4.06 (1.1-14.97; 0.035) | 4.22 (1.47-12.12; 0.008) | 0.74 (0.40-1.38; 0.341) | 0.88 (0.47-1.64; 0.679) | 1.49 (0.90-2.49; 0.123) | 1.46 (0.95-2.25; 0.087) |
| Median income level (50,000-89,999) | | | | | | |
| <50,000 | 1.86 (0.65-5.29; 0.248) | 1.10 (0.48-2.55; 0.822) | 1.42 (1.06-1.91; 0.019) | 1.39 (1.03-1.86; 0.029) | 1.03 (0.78-1.35; 0.853) | 1.04 (0.78-1.38; 0.781) |
| 90,000-129,999 | 1.05 (0.41-2.69; 0.921) | 0.62 (0.27-1.42; 0.255) | 1.03 (0.79-1.33; 0.851) | 0.85 (0.65-1.11; 0.239) | 0.80 (0.62-1.04; 0.100) | 0.63 (0.46-0.85; 0.003) |
| ≥130,000 | 2.05 (0.56-7.50; 0.278) | 1.33 (0.45-3.88; 0.604) | 1.25 (0.80-1.95; 0.327) | 1.12 (0.70-1.79; 0.628) | 0.92 (0.59-1.44; 0.730) | 0.97 (0.63-1.48; 0.875) |
| Tobacco use (Never) | | | | | | |
| Yes | 1.08 (0.35-3.28; 0.892) | 0.65 (0.28-1.5; 0.315) | 1.42 (1.06-1.89; 0.017) | 1.56 (1.17-2.09; 0.003) | 1.21 (0.91-1.6; 0.191) | 1.27 (0.93-1.72; 0.133) |
| Quitted | 1.08 (0.41-2.82; 0.883) | 0.84 (0.39-1.83; 0.666) | 0.84 (0.63-1.12; 0.225) | 1.09 (0.83-1.44; 0.522) | 0.96 (0.73-1.26; 0.782) | 0.98 (0.75-1.29; 0.899) |
| Other | 0.00 (0.00-Inf; 1.00) | 0.00 (0.00-0.00; <0.001) | 10.55 (1.38-80.41; 0.023) | 10.97 (6.25-19.25; <0.001) | 0.00 (0.00-Inf; 0.991) | 0.00 (0.00-0.00; <0.001) |
| Alcohol use (No) | | | | | | |
| Yes | 1.07 (0.39-2.91; 0.899) | 0.81 (0.37-1.78; 0.597) | 0.86 (0.65-1.13; 0.266) | 0.78 (0.58-1.04; 0.085) | 1.22 (0.94-1.59; 0.140) | 1.25 (0.95-1.66; 0.113) |
| Not currently | 1.53 (0.62-3.79; 0.358) | 0.99 (0.44-2.20; 0.979) | 1.10 (0.85-1.43; 0.482) | 1.06 (0.81-1.39; 0.647) | 1.06 (0.82-1.37; 0.660) | 1.09 (0.81-1.46; 0.569) |
| Language (English) | | | | | | |
| Spanish | 0.00 (0.00-Inf; 0.999) | 0.00 (0.00-0.00; <0.001) | 1.13 (0.48-2.65; 0.772) | 1.00 (0.38-2.59; 0.994) | 1.85 (0.96-3.59; 0.067) | 2.6 (1.56-4.33; <0.001) |
| Other | 0.56 (0.00-Inf; 1.00) | 5.17 (1.4-19.03; 0.014) | 0.14 (0.02-1.18; 0.070) | 0.44 (0.05-3.96; 0.462) | 0.76 (0.09-6.21; 0.796) | 0.78 (0.33-1.84; 0.578) |
| Depression (No) | | | | | | |
| Yes | 1.19 (0.52-2.69; 0.682) | 1.25 (0.64-2.41; 0.515) | 1.09 (0.85-1.40; 0.480) | 1.12 (0.86-1.46; 0.393) | 1.12 (0.88-1.43; 0.368) | 1.16 (0.89-1.52; 0.260) |
| Anxiety (No) | | | | | | |
| Yes | 1.16 (0.47-2.88; 0.746) | 1.36 (0.63-2.95; 0.436) | 0.71 (0.54-0.95; 0.022) | 0.84 (0.64-1.11; 0.222) | 1.11 (0.85-1.46; 0.435) | 1.20 (0.88-1.64; 0.252) |
| Other mood disorder (No) | | | | | | |
| Yes | 0.43 (0.15-1.21; 0.109) | 0.69 (0.31-1.57; 0.380) | 0.91 (0.71-1.18; 0.493) | 0.89 (0.66-1.18; 0.409) | 1.36 (1.07-1.73; 0.011) | 1.37 (1.07-1.77; 0.014) |
| Alcohol use disorder (No) | | | | | | |
| Yes | 1.64 (0.50-5.36; 0.412) | 1.12 (0.41-3.10; 0.826) | 1.35 (0.90-2.02; 0.147) | 1.48 (0.86-2.56; 0.160) | 1.02 (0.68-1.51; 0.941) | 1.23 (0.75-2.00; 0.409) |
| Opioid use disorder (No) | | | | | | |
| Yes | 4.84 (1.11-21.10; 0.036) | 2.95 (0.84-10.45; 0.093) | 1.40 (0.84-2.32; 0.193) | 1.36 (0.77-2.39; 0.288) | 0.94 (0.58-1.50; 0.785) | 1.32 (0.84-2.10; 0.232) |
| Cannabis use disorder (No) | | | | | | |
| Yes | 0.00 (0.00-Inf; 0.998) | 0.00 (0.00-0.00; <0.001) | 0.77 (0.39-1.50; 0.441) | 0.56 (0.27-1.15; 0.115) | 1.28 (0.75-2.18; 0.368) | 1.16 (0.61-2.18; 0.653) |
| Other substance use disorder (No) | | | | | | |
| Yes | 0.97 (0.28-3.38; 0.967) | 2.39 (0.79-7.19; 0.121) | 0.65 (0.43-0.99; 0.043) | 0.92 (0.58-1.46; 0.736) | 1.40 (0.96-2.04; 0.081) | 1.18 (0.78-1.78; 0.442) |
| Psychosis (No) | | | | | | |
| Yes | 1.49 (0.39-5.77; 0.561) | 0.97 (0.31-2.98; 0.952) | 1.04 (0.74-1.45; 0.835) | 0.95 (0.65-1.40; 0.804) | 0.93 (0.68-1.28; 0.650) | 0.88 (0.64-1.21; 0.427) |
| Suicidal behavior (No) | | | | | | |
| Yes | 2.67 (0.72-9.93; 0.142) | 1.21 (0.18-8.08; 0.842) | 1.25 (0.81-1.93; 0.306) | 1.31 (0.82-2.09; 0.263) | 1.85 (1.22-2.80; 0.004) | 2.15 (1.36-3.40; 0.001) |

| | | | | | | |
|---|---|---|---|---|---|---|
| Other psychiatric condition (No) | | | | | | |
| Yes | 0.00 (0.00-Inf; 0.998) | 0.00 (0.00-0.00; <0.001) | 1.92 (1.07-3.44; 0.028) | 1.71 (0.92-3.18; 0.089) | 1.27 (0.73-2.20; 0.394) | 1.52 (0.80-2.89; 0.205) |
| CCI (None) | | | | | | |
| 1-2 | 2.43 (0.80-7.43; 0.118) | 2.08 (0.78-5.60; 0.145) | 1.46 (1.13-1.89; 0.004) | 1.45 (1.11-1.89; 0.006) | 1.01 (0.78-1.29; 0.966) | 0.98 (0.74-1.29; 0.861) |
| 3-4 | 3.70 (0.95-14.41; 0.059) | 1.87 (0.64-5.43; 0.253) | 1.60 (1.13-2.28; 0.009) | 1.52 (1.11-2.09; 0.010) | 1.03 (0.73-1.45; 0.852) | 1.00 (0.69-1.46; 0.984) |
| 5+ | 9.84 (3.27-29.66; <0.001) | 5.74 (2.01-16.41; 0.001) | 2.99 (2.13-4.21; <0.001) | 2.04 (1.39-3.00; <0.001) | 1.43 (1.01-2.02; 0.045) | 1.25 (0.86-1.80; 0.238) |

**Note**: Data are from 2022-2023 Yale-New Haven Health (YNHH). HR, hazard ratio; CCI, Charlson Comorbidity Index. a) not estimated due to convergence issues.
